# Right Ventricular Contractile Reserve: A Scoping Review

**DOI:** 10.1101/2025.05.20.25328005

**Authors:** Peter Oro, Roshan Patel, Amish Jain, Azka Khan, Neil Nero, Bo Xu, Adriano R. Tonelli, Matthew T. Siuba

## Abstract

**Background:** Right ventricular contractile reserve (RVCR) describes the ability of the RV to augment contractility under stress, providing valuable insight into latent RV dysfunction across diverse cardiorespiratory diseases. Despite emerging data, RVCR lacks standardized measurement protocols and clinical definitions.

**Objective:** To systematically describe and assess RVCR assessment and clinical implications across multiple conditions.

**Methods:** We conducted a scoping review of studies evaluating RVCR via invasive or non-invasive modalities during exercise or pharmacologic stress in adult populations. Data extracted included methodology, definitions, RV parameters, and clinical implications.

**Results:** We included 62 studies reporting on 4,024 patients, covering healthy individuals, athletes, pulmonary hypertension (PH), structural heart disease, left ventricular assist device (LVAD) recipients, and other conditions (e.g. amyloidosis and COPD). RVCR was assessed via stress echocardiography, invasive pressure-volume (PV) loops, and cardiac MRI. Healthy populations exhibited robust RVCR, whereas patients with PH, heart failure, LVADs, and cardiomyopathies consistently showed impaired RVCR, correlating with exercise intolerance and adverse clinical outcomes.

**Conclusion:** RVCR is a significant marker of right-heart adaptability across conditions. Heterogeneity in definitions, testing modalities, and imaging techniques limits clinical applicability and generalizability. Standardized methods and prospective validation are needed.

## Introduction

Left ventricular (LV) performance has traditionally been at the forefront of cardiovascular risk stratification. However, even subtle impairments in right ventricular (RV) function can result worse outcomes.^1,2^ RV dysfunction is associated with poor outcomes across a variety of cardiorespiratory disease states, including acute respiratory distress syndrome (ARDS), heart failure (HF), post-cardiac surgery, acute myocardial infarction (MI), congenital heart disease, and pulmonary hypertension (PH).^3–6^ RV function has historically been underappreciated greatly due to its difficult assessment in the context of complex geometry and coupling with the pulmonary circulation.^7^

When required, a normal RV increases its contractility to sustain cardiorespiratory function. Measuring the RV’s capacity to augment contractility in response to physiologic stress is referred to as right ventricular contractile reserve (RVCR). Latent RV dysfunction can be unmasked under stress by suboptimal increase in its contractility. Invasive pressure-volume loop catheterization is considered the gold standard for assessing RVCR. However, other more widely available modalities have been increasingly studied in recent years, including stress right heart catheterization (RHC), non-invasive stress echocardiography and stress cardiac magnetic resonance imaging (CMR).^8,9^ These modalities have been studied across a broad spectrum of conditions, ranging from endurance athletes to patients with HF.

RVCR has not been previously reviewed in a structured fashion. Therefore, we performed a scoping review to characterize RVCR and its clinical implications across a broad spectrum of conditions, including healthy individuals, athletes, PH, structural/functional heart diseases, LVAD, and others.

## 2. Methods

### 2.1. Protocol

A scoping review strategy was chosen due to expected limited availability of large randomized controlled trials or observational studies assessing RVCR. Ethics approval was not required. This review follows the Preferred Reporting Items for Systematic Reviews and Meta-Analyses extension for Scoping Reviews (PRISMA-ScR) guidelines.

### 2.2. Eligibility Criteria

We included randomized controlled trials, retrospective/prospective cohort studies, and non-controlled experimental studies assessing RVCR in adult patients using invasive hemodynamics (e.g. right heart catheterization, PV loops) or non-invasive imaging (e.g. echocardiography, CMR) during exercise testing (e.g. treadmill, bicycle, handgrip) or pharmacologic stress testing (e.g. dobutamine). We excluded case reports, case series, review articles, conference abstracts, and studies without quantitative pre- and post-test hemodynamic or imaging data.

### 2.3. Information Sources and Search Strategy

A systematic search was conducted across MEDLINE, Embase, Cochrane Library, Scopus, Web of Science, and CINAHL from 1979 until July 11, 2024. The search strategy included permutations of RV function, contractile reserve, exercise testing, pharmacologic stress, echocardiography, MRI, and catheterization (see **Online Supplement**). Studies were restricted to adult humans.

### 2.4. Selection of Sources of Evidence

We included studies that reported RVCR measurements before and after stress and excluded those lacking quantitative data or specific stress modalities. Two independent reviewers screened abstracts, reviewed full texts, and extracted data, with conflicts resolved by a third reviewer.

### 2.5. Data Charting Process and Data Items

Covidence software (Covidence systematic review software, Veritas Health Innovation, Melbourne, Australia) was used for abstract screening, full text review, and data extraction. Two independent reviewers extracted study details, including study design, patient population, stressor type (exercise vs. pharmacologic), and imaging/hemodynamic modalities (echocardiography, MRI, right heart catheterization, PV loops). We recorded pre- and post-test RV parameters from various imaging modalities, as well as the definition and magnitude of RVCR and any prognostic outcomes. A third reviewer verified all data for accuracy.

### 2.6. Synthesis of Results

No formal synthesis was performed based on the scoping design.

## 3. Results

After removal of duplicates, 436 studies were screened. A total of 177 studies were assessed in full text review, and ultimately 62 studies including 4,024 patients were included. Details of screening are included in **Figure 1**. A summary of major findings is included below, as well as **Tables 1-5** and **Figure 2**.

**Figure 1.**
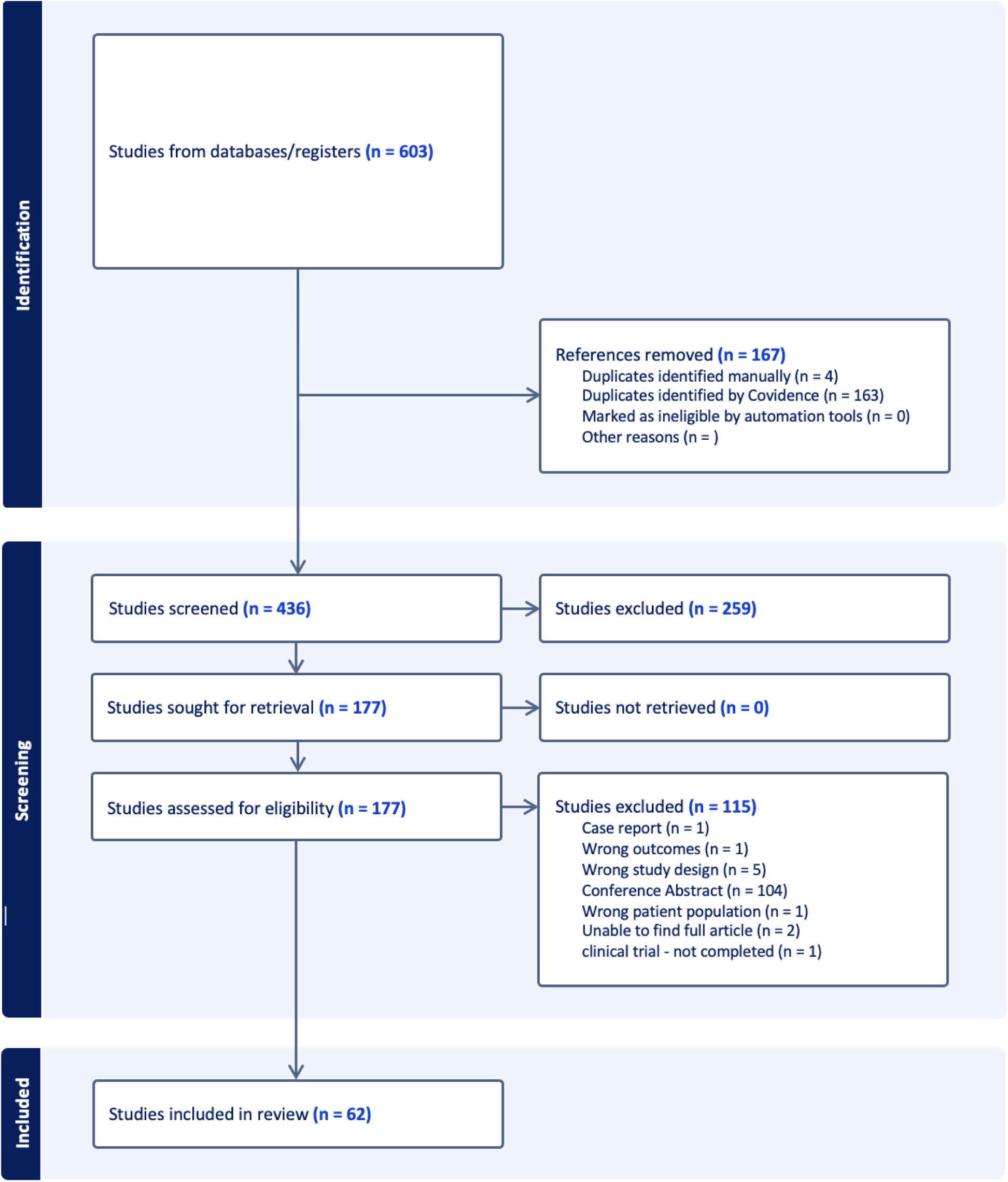
CONSORT diagram of included studies

**Figure 2.**
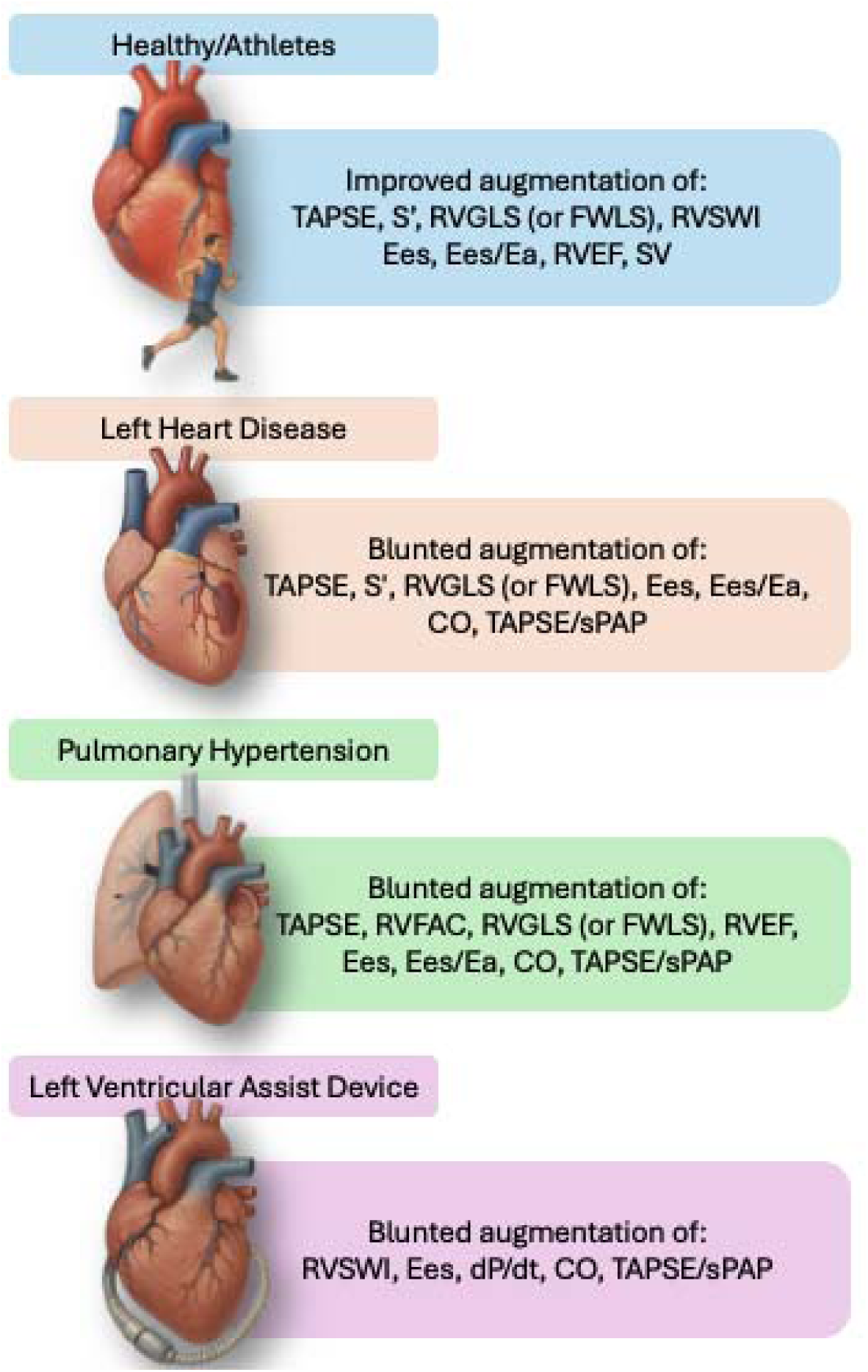
Summary of Commonly Assessed RVCR Parameters with Stress Across Various Conditions. Note: “Blunted augmentation” indicates reduced stress-induced change compared to healthy individuals or controls. Abbreviations: CO = cardiac output; dP/dt = rate of pressure rise; Ea = arterial elastance; Ees = end-systolic elastance; FWLS = free wall longitudinal strain; RVEF = right ventricular ejection fraction; RVGLS = right ventricular global longitudinal strain; RVSWI = right ventricular stroke work index; S′ = systolic annular velocity; sPAP = systolic pulmonary artery pressure; TAPSE = tricuspid annular plane systolic excursion.

**Table 1:**
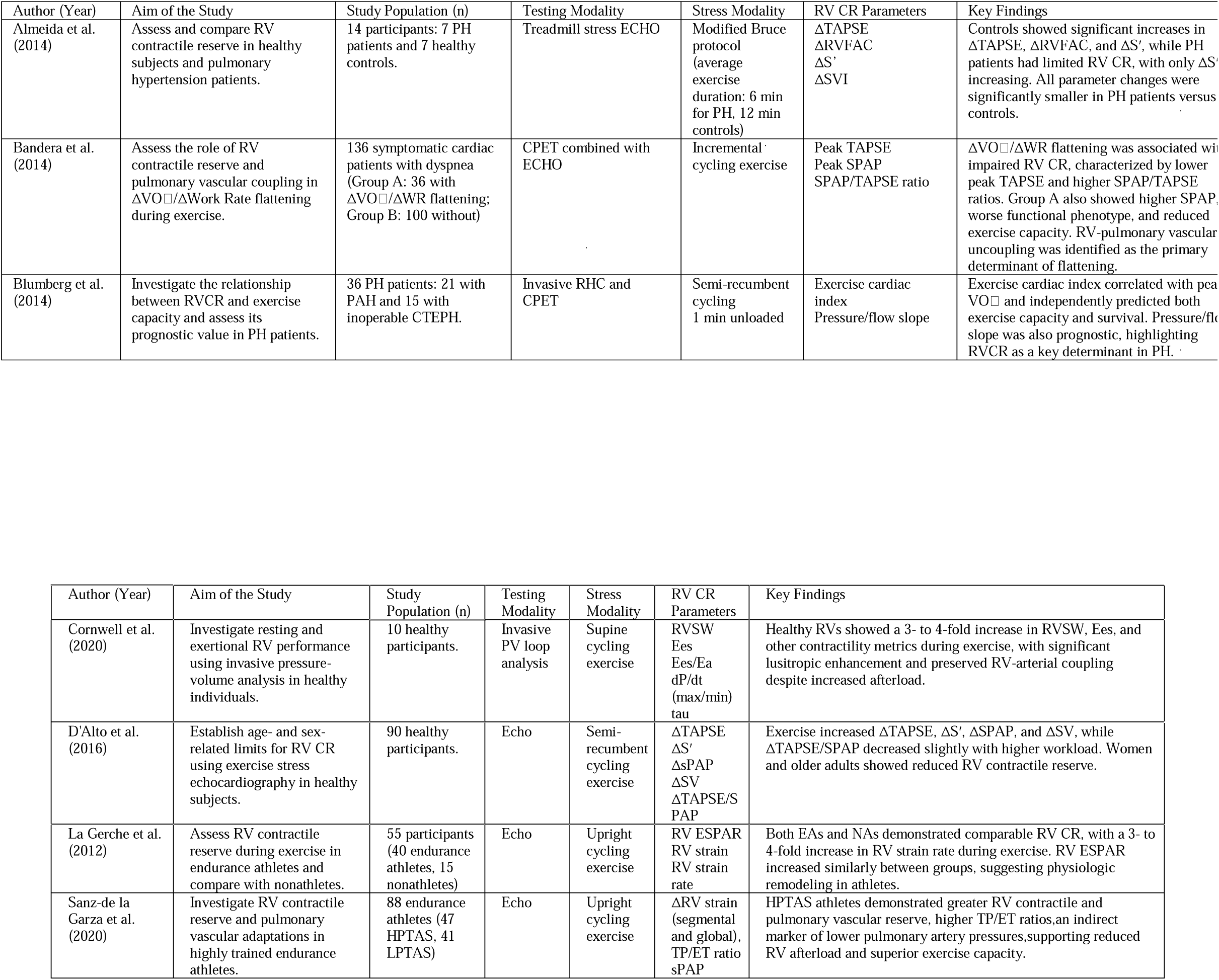
Summary of Studies Assessing Right Ventricular Contractile Reserve in Healthy and Athletic Participants. RV = Right Ventricle; CR = Contractile Reserve; PV = Pressure-Volume; RVSW = Right Ventricular Stroke Work; Ees = End-Systolic Elastance; Ees/Ea = RV-Pulmonary Arterial Coupling; dP/dt (max/min) = Maximum/Minimum Rate of Pressure Change; tau = Time Constant of Isovolumic Relaxation; TAPSE = Tricuspid Annular Plane Systolic Excursion; S′ = Systolic Annular Velocity; SPAP = Pulmonary Artery Systolic Pressure; SV = Stroke Volume; TAPSE/sPAP = Ratio of TAPSE to sPAP; RV ESPAR = Right Ventricular End-Systolic Pressure-Area Relationship; RV SR = Right Ventricular Strain Rate; ΔRV strain = Change in Right Ventricular Strain; TP/ET ratio = Tricuspid Annular Plane to Ejection Time Ratio; EAs = Endurance Athletes; NAs = Nonathletes; HPTAS = High Pulmonary Training Adaptation Status; LPTAS = Low Pulmonary Training Adaptation Status.

### 3.1 Healthy Individuals & Athletes

Four studies, including 243 participants, evaluated RVCR in healthy individuals (n = 115) and athletes (n = 128). These studies aimed to identify normal RVCR, comparing athletes and other healthy individuals or different groups of athletes. The most common method of assessing RVCR was exercise stress echocardiography (three studies), with one using PV loops. Two of the three echocardiography studies utilized upright cycling, while the remaining utilized semi-recumbent cycling, and the PV study utilized supine cycling. A detailed summary of these studies, including population characteristics, assessment modalities, and key findings, is provided in **Table 1**.

Using invasive PV loops in healthy individuals, Cornwell et al. found that individuals with healthy RVs had robust RVCR during exercise, noting a three- to four-fold increase in RV stroke work (RVSW) (1,605±663 to 4,999±2,342 mmHg*ml) and end-systolic elastance (Ees) (0.3±0.2 to 0.9±0.8 mmHg/ml).^8^ They also demonstrated significant improvement in RV diastolic function (lusitropic reserve) during exercise, characterized by a reduction in Tau (49 ± 12 msec at rest to 33 ± 8 msec at peak exercise) and an increase in dP/dt min, reflecting enhanced active relaxation under stress conditions. The RV-PA coupling also improved (Ees/arterial elastance [Ea]) (1.2±0.5 at rest and 1.6±0.8 with peak exercise), despite the increased afterload observed.

From the echocardiographic studies, D’Alto et al. found that healthy individuals experienced significant exercise-induced increases in tricuspid annular plane systolic excursion (TAPSE) from 27±2 to 34±2 mm, tissue Doppler velocity of the tricuspid annulus (S′) from 15±2 to 25±4 cm/s, pulmonary artery systolic pressure (sPAP) from 22±5 to 34±7 mm Hg, and stroke volume (SV) from 71±17 to 120±28 mL.^10^ Women and older subjects had lower RVCR when compared to men and younger participants, and the maximum exercise capacity directly correlated with the change in TAPSE/sPAP. La Gerche et al. compared endurance athletes to non-athletes and found that both groups had comparable RVCR as assessed by the RV end-systolic pressure-area ratio (RVESPAR).^11^ RVESPAR, calculated as sPAP divided by RV end-systolic area, increased in both groups, from 1.3 at rest to 10.0 mmHg/cm² at peak exercise in endurance athletes, and from 1.5 mmHg/cm² at rest to 9.3 mmHg/cm² at peak exercise in nonathletes. These findings support the interpretation that preserved RV functional reserve, despite reduced resting RV strain in endurance athletes, likely reflect physiologic remodeling rather than subclinical dysfunction. Finally, Sanz-de la Garza et al. compared endurance athletes with high versus low pulmonary transit of agitated saline (PTAS), measured by contrast echocardiography during exercise as the number of bubbles appearing in the left atrium after venous injection, a surrogate of pulmonary vascular reserve. Athletes with high PTAS (>12 bubbles) exhibited greater RVCR as reflected by higher peak TAPSE (33.0 ± 4.2 vs. 30.9 ± 3.5 mm), and enhanced pulmonary vascular reserve, reflected by a higher time-to-peak/ejection time (TP/ET) ratio (46.9±4.6 vs. 43.3±5.6), derived from right ventricle outflow tract Doppler signals. These improvement in RV contractile and pulmonary vascular reserve parameters correlated with superior exercise capacity, including higher VO max (∼65 vs. ∼55 mL/kg/min) and workload (∼370 vs. ∼330 W).^12^

### 3.2 Pulmonary Hypertension (PH)

A total of 24 studies including 1,390 patients evaluated RVCR across multiple PH subgroups and associated conditions, such as systemic sclerosis (SSc), idiopathic pulmonary fibrosis (IPF), obesity, and pulmonary valve stenosis (PS). Various testing modalities were utilized to measure RVCR, including echocardiography (8 studies), RHC (7 studies), invasive PV loops (5 studies), CMR (4 studies), and speckle-tracking MRI (1 study). Different interventions were used as stressors, including various cycling protocols (incremental, supine, semi-supine, upright) in 12 studies, treadmill testing (graded, modified Bruce protocol) in 2 studies, dobutamine stress echocardiography in 2 studies, and leg-positive pressure maneuver in one study. A detailed summary of these studies, including methodology, patient populations, and key findings, is provided in **Table 2**.

**Table 2:**
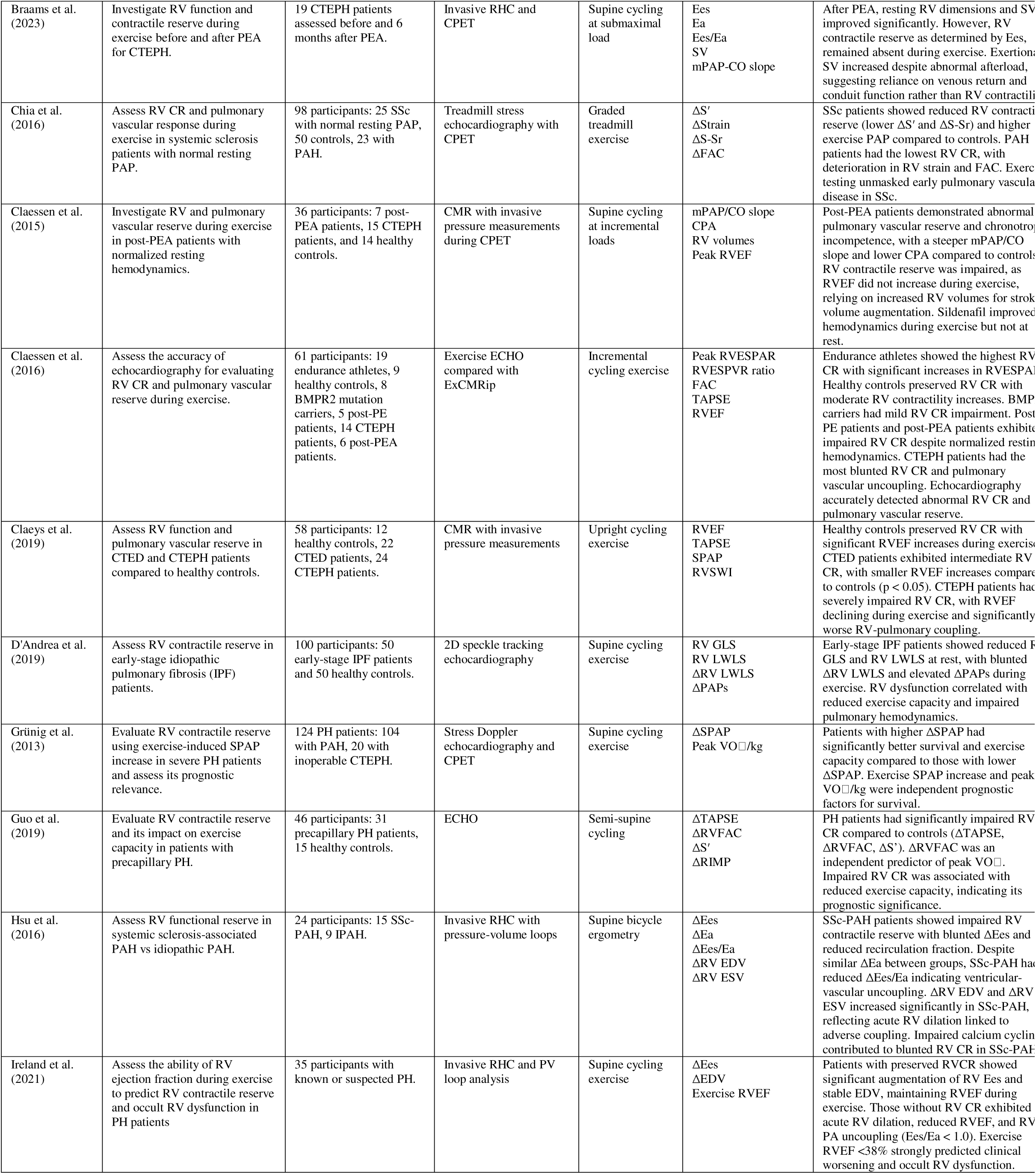
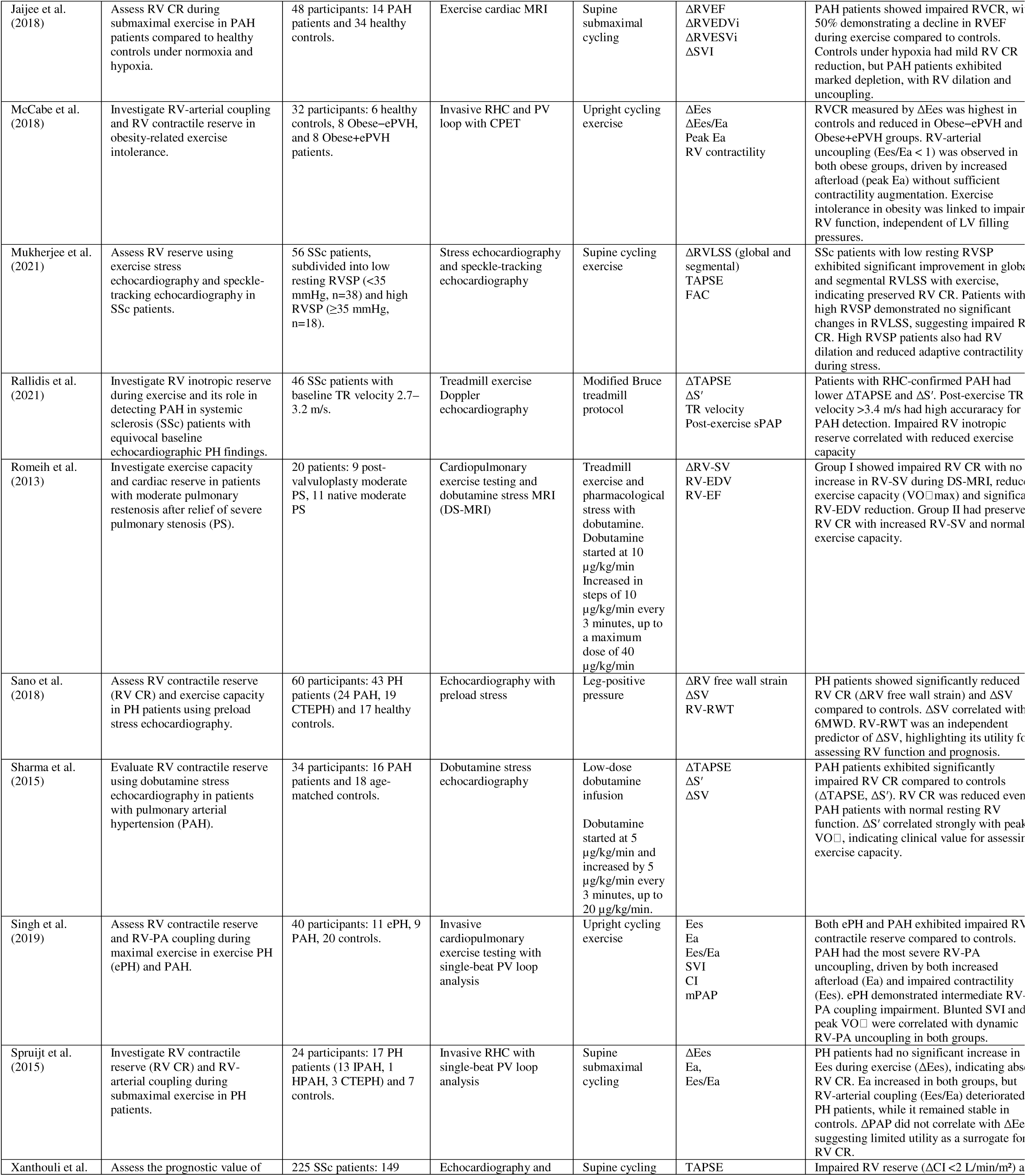
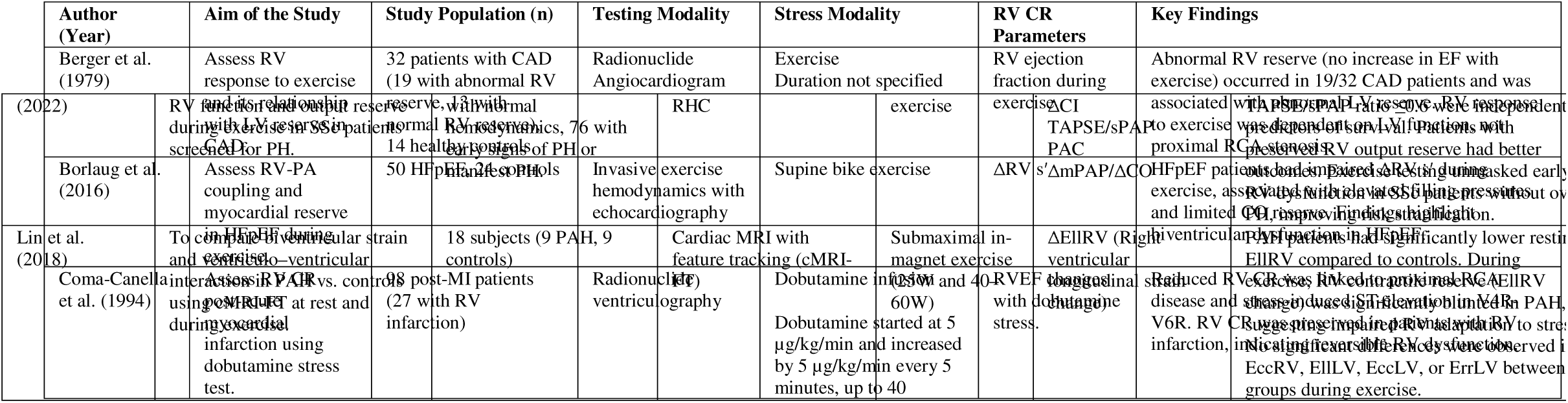
Summary of Studies Assessing Right Ventricular Contractile Reserve in Pulmonary Hypertension and Related Conditions. RV = Right Ventricle; CR = Contractile Reserve; CPET = Cardiopulmonary Exercise Testing; ECHO = Echocardiography; CMR = Cardiac Magnetic Resonance Imaging; TDI = Tissue Doppler Imaging; HF = Heart Failure; HFpEF = Heart Failure with Preserved Ejection Fraction; HTDD = Hypertensive Type Diastolic Dysfunction; CTEPH = Chronic Thromboembolic Pulmonary Hypertension; CMS = Chronic Mountain Sickness; HA = High-Altitude Dwellers; ID = Iron Deficiency; COPD = Chronic Obstructive Pulmonary Disease; ATTRwt-CA = Wild-type Transthyretin Cardiac Amyloidosis; T1D = Type 1 Diabetes; NAC = National Amyloidosis Centre; TAPSE = Tricuspid Annular Plane Systolic Excursion; RVESPAR = Right Ventricular End-Systolic Pressure–Area Relationship; RVFWSRL = Right Ventricular Free Wall Strain Rate Longitudinal; RVGLS = Right Ventricular Global Longitudinal Strain; RVLS = Right Ventricular Longitudinal Strain; FAC = Fractional Area Change; S′ = Peak Systolic Velocity of the Tricuspid Annulus (Tissue Doppler); ΔRVGLS = Change in Right Ventricular Global Longitudinal Strain; ΔRV-S′ = Change in Right Ventricular S′ with Stress; ΔRV FAC = Change in Right Ventricular Fractional Area Change; SPAP = Pulmonary Artery Systolic Pressure; sPAP = Systolic Pulmonary Arterial Pressure; SV = Stroke Volume; SVI = Stroke Volume Index; ΔEF = Change in Ejection Fraction; ΔGLS = Change in Global Longitudinal Strain; ΔGCS = Change in Global Circumferential Strain; GRS = Global Radial Strain; ΔRVSWI = Change in Right Ventricular Stroke Work Index; ΔPAWP/ΔCO = Change in Pulmonary Artery Wedge Pressure per Change in Cardiac Output; VO[ = Oxygen Consumption; LVEF = Left Ventricular Ejection Fraction; LVGLS = Left Ventricular Global Longitudinal Strain; LA = Left Atrium; TR = Tricuspid Regurgitation; EccRV = Right Ventricular Circumferential Strain; EllLV = Left Ventricular Longitudinal Strain; EccLV = Left Ventricular Circumferential Strain; ErrLV = Left Ventricular Radial Strain.

In a study of patients with PAH and inoperable CTEPH, low exercise cardiac index, used as a surrogate for RVCR, correlated with reduced peak VO. Cardiac index increased from 2.2±0.6 to 3.4±1.1 L/min/m², a blunted rise (∼1.2 L/min/m²) compared to the ≥2.0 L/min/m² increase of healthy individuals. This impaired RVCR was the only independent predictor of peak VO and a significant prognostic marker.^13^ Similarly, an increase in sPAP of >30 mmHg with exercise, measured via stress echocardiography and used as a surrogate for preserved RVCR, was independently associated with better survival in patients with PAH and inoperable CTEPH.^14^ This aligns with findings from Sano et al., where preload stress was achieved through a leg-positive pressure maneuver increasing venous return. There was significantly lower ΔRV free wall strain (8.2±11.9% vs. 14.5±6.6% in controls) and ΔSV (3.6±6.0 mL vs. 8.5±2.3 mL in controls) in PH patients (PAH and CTEPH) compared to controls, indicating RVCR impairment even under conditions of isolated preload augmentation.^15^ Invasive PV loop analysis demonstrated that PH patients (including idiopathic and hereditary PAH as well as CTEPH) failed to have a significant increase in Ees during exercise, in contrast to healthy controls.^16^

Several other studies demonstrated that PAH patients exhibit impaired RVCR under stress. Almeida et al. found that PAH patients had significantly lower exercise-induced increases in TAPSE and RVFAC during treadmill stress echocardiography compared to healthy controls. TAPSE increased only modestly in PH patients (from 16.1±4.6 to 16.8±2.7 mm) versus a marked increase in controls (from 25.7±2.4 to 31.0±3.5 mm), and RVFAC remained unchanged in PH patients (19.8±9.7 to 19.4±14.4%) while it increased in controls (53.8±14.7 to 64.4±9.9%).^17^ Similarly, semi-supine cycling stress echocardiography revealed impaired RVCR in PH patients, with TAPSE increasing from 17.5±3.5 to 18.3±2.6 mm and RVFAC from 33.4±7.5 to 36.6±8.6%, compared to healthy controls who increased TAPSE from 22.5±2.6 to 28.9±2.6 mm and RVFAC from 43.8±3.8 to 55.1±2.6%.^18^ Impairment was also evident during dobutamine stress echocardiography, where ΔTAPSE and ΔSV were significantly reduced.^19^ Using exercise CMR, Jaijee et al. showed that at least 50% of PAH patients exhibited a decline in RVEF during supine submaximal cycling despite preserved resting function (PAH: rest 53.6±4.3% vs. exercise 51.4±10.7%; controls: rest 57.1±5.2% vs. exercise 69.6±6.1%).^20^

RVCR and RV-PA coupling were also assessed using invasive cardiopulmonary exercise testing (CPET) with PV loop analysis in PAH and exercise PH patients.^21^ PAH patients had the most severe RV–PA uncoupling, primarily driven by markedly elevated afterload (peak Ea 1.84±0.46 mmHg/mL/m²) and impaired contractile reserve (ΔEes 0.41±0.87 mmHg/mL/m²), resulting in the lowest peak RV–PA coupling ratio (Ees/Ea 1.06±0.38). Similarly, Ireland et al. demonstrated that patients with impaired RVCR exhibited acute RV dilation and reduced exercise RVEF. An exercise RVEF <38% predicted occult RV-PA uncoupling (Ees/Ea <1.0) with 85% sensitivity and 77% specificity, and was associated with a higher risk of clinical worsening over 4 years.^22^

In SSc patients with normal resting pulmonary artery pressure, treadmill stress echocardiography unmasked early pulmonary vascular dysfunction. SSc patients had reduced S′ both at rest (10.2±2.1 vs. 11.6±1.0 cm/s) and post-exercise (16.4±3.0 vs. 19.5±2.3 cm/s), compared to controls. Systolic PAP rose to higher levels in SSc despite similar resting values (rest: 27.8±5.4 vs. 22.7±6.9 mmHg; post-exercise: 42.8±11.5 vs. 29.3±11.8 mmHg). In PAH patients, S′ increased from 9.9±1.9 to 12.4±3.9 cm/s and sPAP from 68.6±32.0 to 83.0±30.0 mmHg.^23^ This findings were reinforced by Mukherjee et al., who found that SSc patients with low resting RVSP (<35 mmHg) exhibited preserved RVCR, evidenced by a significant increase in global RV longitudinal systolic strain (RVLSS) from −15.3±3.5% at rest to −19.0±4.5% with exercise. In contrast, patients with elevated RVSP (≥35 mmHg) at rest showed no significant change in RVLSS (−17.6±4.2% at rest vs. −18.7±3.8%), and developed RV dilation with exercise.^24^ Rallidis et al. showed that smaller changes in TAPSE (ΔTAPSE 3.4±2.0 mm in PAH vs. 5.4±3.0 mm in non-PAH) and S′ (ΔS′: 4.3±3.2 cm/s vs. 6.3±3.1 cm/s), helped distinguish SSc patients with PAH from those without. Post-exercise sPAP was significantly elevated in the PAH group (61.6 ±11.0 mmHg) compared to the non-PAH group (42.7±7.1 mmHg).^25^ In SSc-associated PAH, invasive hemodynamic studies showed a lack of increase in RV contractility (measured by Ees) during exercise, in contrast to idiopathic PAH patients, who showed preserved RV contractile reserve (Ees increase from 1.23±0.15 to 2.32±0.28 mmHg/mL). To compensate, SSc-PAH patients exhibited acute RV dilation, with RV end-diastolic volume increasing by 37±10% and end-systolic volume by 19±4%, while idiopathic PAH patients maintained stable volumes.^26^ Complementing this, Xanthouli et al. showed that impaired RV output reserve, defined as a cardiac index increase during exercise of <2 L/min/m², and a low TAPSE/sPAP ratio (≤0.6 mm/mmHg) were predictors of mortality in a large cohort of SSc patients undergoing PH screening.^27^

Both CTEPH and CTED patients demonstrate impaired RVCR during exercise. Exercise CMR coupled with invasive pressure measurements revealed that CTEPH patients had severely blunted RVCR, quantified by a peak-to-rest RV end-systolic pressure–volume ratio (RVESPVR) of 1.35±0.23, and a decline in RVEF from 36.8±9.7 to 33.3±9.1%. CTED patients exhibited intermediate impairment, with RVESPVR of 2.23±0.55 and a modest increase in RVEF from 57.8±6.5 to 62.1±8.9%, suggesting subclinical dysfunction unmasked only with exertion.^28^ Claessen et al. found that patients with CTEPH, CTEPH post-PTE (pulmonary thromboendarterectomy), and patients post-PE (pulmonary embolism) exhibited impaired RVCR during exercise, with similarly reduced RVESPVR ratios in CTEPH (1.45±0.30) and post-PTE patients (1.45±0.38), while post-PE patients showed abnormal pulmonary vascular reserve with less RVCR impairment.^29^ An earlier study revealed that CTEPH patients who underwent PTE patients normalized resting hemodynamics, but failed to augment RV contractility with exercise and relied on RV dilation to maintain SV.^30^ Braams et al. extended these findings, showing that although SV increased with exercise after PTE (79±32 to 102±28 mL), RV Ees remained roughly unchanged (rest: 0.32[0.23–0.40] vs exercise: 0.28[0.19–0.44] mmHg/mL).^31^

In early IPF, D’Andrea et al. demonstrated significantly reduced exercise-induced changes in RV global strain (ΔRV GLS: 5.9±2.4% in IPF vs. 8.2±2.6% in controls) and RV lateral wall longitudinal strain (ΔRV LWLS: 22.2±0.9% vs. 31.3±2.2%). These reduction in strain correlated with elevated pulmonary pressures and reduced exercise capacity.^32^ McCabe et al. demonstrated impaired RV contractile reserve in obese patients, both without (Obese–ePVH) and with (Obese+ePVH) exercise pulmonary venous hypertension (ePVH). ΔEes increased only to 154 ± 39% of baseline in Obese–ePVH and 141 ± 34% in Obese+ePVH, compared to 224 ± 80% in healthy controls. Although peak RV Ees was similar across groups (controls: 0.50 ± 0.16 mmHg/mL; Obese–ePVH: 0.45 ± 0.22 mmHg/mL; Obese+ePVH: 0.48 ± 0.17 mmHg/mL), obese patients exhibited significantly higher afterload (peak Ea: 0.31 ± 0.07 mmHg/mL in controls vs. 0.75 ± 0.32 and 0.88 ± 0.62 mmHg/mL in Obese–ePVH and Obese+ePVH, respectively), contributing to reduced exercise capacity.^33^ Bandera et al. demonstrated that patients with flattening of the ΔVO /ΔWR slope, reflecting impaired oxygen uptake relative to work rate, had reduced RV contractile reserve, shown by lower peak TAPSE (22±5 vs. 25±7 mm), and greater RV-pulmonary vascular uncoupling, reflected by a higher sPAP/TAPSE ratio (3.03±1.25 vs. 2.29±1.34), compared to patients with a normal ΔVO /ΔWR response.^34^ Lin et al. used CMR feature tracking and found that patients with PAH exhibited blunted RV longitudinal strain augmentation (during in-magnet submaximal cycling compared to controls (Δ–1.1±3.7 vs –3.9±3.4%), despite similar resting RVEF.^35^ Similarly, RVCR was diminished in patients with moderate pulmonary valve restenosis after prior balloon valvuloplasty or surgical valvotomy for severe pulmonary valve stenosis. Romeih et al. demonstrated a decrease in RV stroke volume during dobutamine stress (ΔSV: – 5 ± 8 mL vs. +13 ± 8 mL in controls), and this impairment correlated with reduced exercise capacity, as evidenced by a significantly lower peak VO (72.8 ± 3.5% vs. 102.5 ± 16.3% of predicted). ^36^

### 3.3 Structural and Functional Heart Diseases

A total of 20 studies including 1,476 patients evaluated RVCR in patients with coronary artery disease (CAD), heart failure with preserved ejection fraction (HFpEF), heart failure with reduced ejection fraction (HFrEF), hypertrophic cardiomyopathy (HCM), post-myocardial infarction (MI), idiopathic dilated cardiomyopathy (IDC), and mitral valve disease. Fourteen studies assessed RVCR using exercise stress while six studies used pharmacologic stress.

Exercise testing (cycling, treadmill, or handgrip exercise) was combined with stress echocardiography (8 studies), RHC (3 studies), radionuclide ventriculography (2 studies), and invasive PV loop analysis (1 study). Pharmacologic stress testing included dobutamine echocardiography (4 studies) and radionuclide ventriculography (2 studies). A summary of these studies is provided in **Table 3**.

**Table 3:**
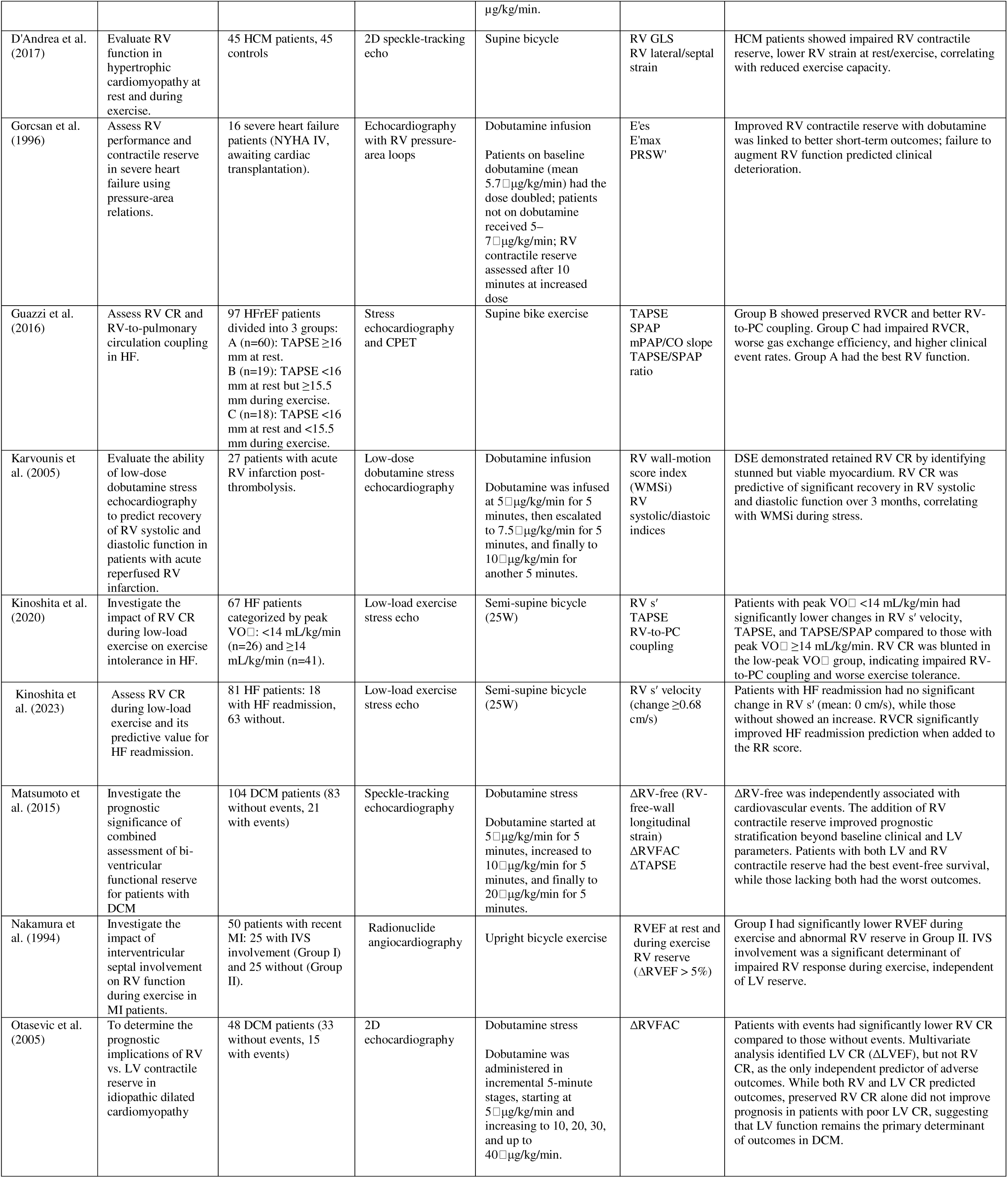
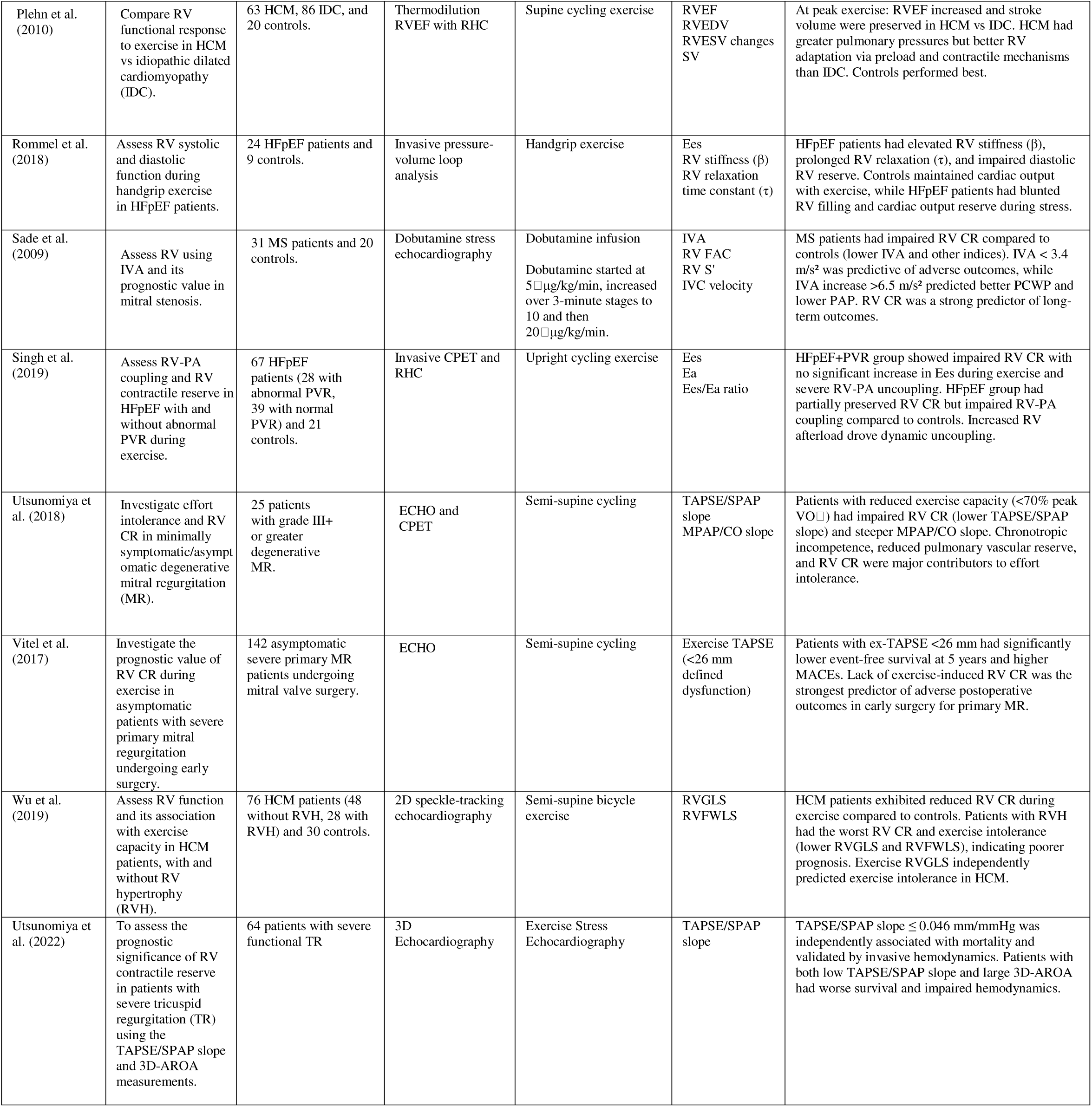
Studies Investigating Right Ventricular Contractile Reserve in Patients with Left Heart Disease. RV = Right Ventricle; CR = Contractile Reserve; ATTRwt-CA = Wild-Type Transthyretin Cardiac Amyloidosis; CAD = Coronary Artery Disease; HF = Heart Failure (HFpEF = Heart Failure with Preserved Ejection Fraction, HFrEF = Heart Failure with Reduced Ejection Fraction); CPET = Cardiopulmonary Exercise Testing; ECHO = Echocardiography; RHC = Right Heart Catheterization; TAPSE = Tricuspid Annular Plane Systolic Excursion; RVGLS = Right Ventricular Global Longitudinal Strain; RVFWLS = Right Ventricular Free-Wall Longitudinal Strain; SPAP = Pulmonary Artery Systolic Pressure; mPAP = Mean Pulmonary Arterial Pressure; CO = Cardiac Output; SPAP = Systolic Pulmonary Artery Pressure; MPAP/CO slope = Slope of the Mean Pulmonary Artery Pressure to Cardiac Output; IVA = Isovolumic Acceleration; FAC = Fractional Area Change; Ees = End-Systolic Elastance (RV Contractility); Ea = Effective Arterial Elastance (Pulmonary Arterial Load); Ees/Ea = RV-PA Coupling (Right Ventricle to Pulmonary Artery Coupling); PRSW = Preload Recruitable Stroke Work; WMSi = Wall-Motion Score Index; VO□ = Oxygen Consumption; PCWP = Pulmonary Capillary Wedge Pressure; MACEs = Major Adverse Cardiac Events; LV = Left Ventricle; RVEF = Right Ventricular Ejection Fraction; RVEDV = Right Ventricular End-Diastolic Volume; RVESV = Right Ventricular End-Systolic Volume; NYHA = New York Heart Association (Functional Class).

In HFpEF, impaired RV adaptation to exercise was consistently observed across several studies. Borlaug et al. demonstrated that, compared to controls, HFpEF patients exhibited smaller increases in RV S′ (from 10.7±2.4 to 14.0±3.8 cm/s in HFpEF vs. 10.6±2.4 to 17.4±3.7 cm/s in controls) and RV e′ (from 7.6±2.0 to 9.7±2.8 cm/s vs. 8.1±2.1 to 12.1±2.3 cm/s). This was accompanied by lower pulmonary artery compliance at peak exercise (2.0±0.9 mL/mmHg vs. 3.0±1.5 mL/mmHg) and blunted cardiac output (CO) augmentation (9.2±2.1 vs. 11.4±2.4 L/min) when HFpEF patients were compared with controls.^37^ Expanding on these findings, Singh et al. reported that HFpEF patients with abnormal pulmonary vascular resistance (PVR) during upright cycling only increased Ees from 0.98±0.44 to 1.05±0.59 mmHg/mL, compared to 1.38±0.57 to 1.86±0.82 mmHg/mL in those with normal PVR. This resulted in more pronounced RV-PA uncoupling, as reflected by a lower exercise Ees/Ea ratio (0.68±0.25 vs. 1.02±0.41).^38^ Complementing these insights, Rommel et al. identified impaired RV diastolic reserve in HFpEF compared to controls, with increased RV stiffness at rest (0.018±0.003 vs. 0.012±0.003 mmHg/mL) and prolonged relaxation time (τ) during handgrip exercise (+6.8±6.0 vs. –5.3±4.4 ms), contributing to limited CO augmentation and exercise intolerance.^39^

In HFrEF, Guazzi et al. identified three distinct RV phenotypes based on TAPSE responses during exercise. Patients with preserved RV function at rest (group A; TAPSE ≥16 mm) demonstrated better RV-PA coupling (peak TAPSE/PASP ratio: 0.42±0.14 mm/mmHg) and superior exercise tolerance. Among patients with reduced resting TAPSE (<16 mm), two subgroups were distinguished based on exercise performance: Group B with preserved RVCR (peak TAPSE ≥15.5 mm) and Group C with severely impaired RVCR (peak TAPSE <15.5 mm). Compared to Group B, Group C exhibited worse RV-PA uncoupling (peak TAPSE/PASP: 0.21±0.08 vs. 0.34±0.11 mm/mmHg), higher resting PVR (3.5±1.7 vs. 2.8±1.2 WU), and elevated resting mPAP (30.8±12.7 vs. 23.8±9.3 mmHg), along with poorer ventilatory efficiency.^40^ Gorcsan et al. utilized RV pressure-area relations, analogous to PV loops, by combining high-fidelity RHC with echocardiographic automated border detection of RV area during transient IVC balloon occlusion to assess RVCR in patients with severe heart failure. They found that an inability to augment RV Ees and maximal elastance (E’max) in response to dobutamine infusion was associated with poor short-term outcomes, including the need for mechanical circulatory support. Specifically, patients who achieved ≥100% increases in Ees (from 2.9± 1.5 to 5.5±3.3 mmHg/cm²) or E’max (from 3.3±1.6 to 6.4±3.9 mmHg/cm²) were clinically stable, while those unable to augment RV contractility deteriorated.^41^

Kinoshita et al. demonstrated that RVCR, assessed by the change in RV S′ during low-work load (25W) exercise stress echocardiography, strongly predicted HF readmission. Patients with impaired RVCR, defined as a ΔRV S′ ≤0.68 cm/s,had significantly higher HF readmission rates (48.5% at 1 year) compared to those with preserved RVCR (0% readmission).^42^ Similarly, the change in RV S′ velocity at 25W correlated closely with VO_2_. A ΔRV S′ cut-off of 0.68 cm/s effectively identified those with peak VO <14 mL/kg/min, a cut-off associated with 1-year mortality and used to identify candidates that may benefit from heart transplantation.^43^

In HCM, D’Andrea et al. demonstrated reduced right ventricular longitudinal strain (RVGLS) at rest (12.8±3.6 vs. 19.7±3.6% in controls) and during exercise (13.8±4.3 vs. 24.7±3.7%), that was directly associated with lower exercise capacity.^44^ Building on this, Wu et al. showed that HCM patients with RV hypertrophy (RVH) had even greater impairments in RV function, with less negative strain values: exercise RVGLS (–13.4±2.4%) and RV free wall longitudinal strain (RVFWLS) (–16.7±1.8%) in RVH patients, compared to patients without RVH (RVGLS –17.1±2.3%; RVFWLS –18.8±2.4%) and controls (RVGLS –22.3±1.7%; RVFWLS –25.2±1.2%). These impairments, particularly exercise RVGLS, correlated with lower exercise capacity.^45^ In IDC, Plehn et al. demonstrated that RV response to exercise is significantly impaired compared to both hypertrophic cardiomyopathy (HCM) and controls. Specifically, IDC patients failed to augment RV SV (117±47 to 120±52 mL) or RVEF (35±10% to 37±11%) during supine cycling, while controls and HCM patients showed greater increases.^46^ In IDC patients undergoing dobutamine stress echocardiography Otasevic et al. found that those who experienced cardiovascular events had lower RVCR (ΔRVFAC 8±6%) and LV contractile reserve (ΔLVEF 3±2%) compared to event-free patients (ΔRVFAC 14±5%; ΔLVEF 9±5%).^47^ Complementing this, Matsumoto et al. demonstrated that combined assessment of biventricular contractile reserve provided incremental prognostic value in IDC. LV contractile reserve was defined by global circumferential strain (GCS), a measure of mid-wall myocardial shortening, with preserved LV reserve identified by ΔGCS ≥13.7%, and RV reserve defined by ΔRVFWS ≥16.5%. Patients with preserved reserve in both ventricles had the best event-free survival, while those with a reduction in biventricular reserve had the worst outcomes.^48^

In CAD and post-MI patients, several studies have explored the prevalence and impact of RVCR. Berger et al. demonstrated that abnormal RVCR, defined as failure to increase RVEF by at least 5% during upright bicycle exercise, occurred in 19 of 32 CAD patients. All patients with abnormal RVCR also had impaired LV reserve, and the RV function strongly correlated with changes in LVEF, suggesting that RV dysfunction was primarily driven by LV impairment rather than proximal right coronary artery (RCA) stenosis. ^49^ In contrast, Coma-Canella et al. found that in post-MI patients, RVEF augmentation during dobutamine echocardiography was blunted in those with proximal RCA disease (increase: 8±11%) compared to those with distal RCA disease (17 ± 12%) or no RCA disease (17 ± 10%). This blunted RVCR correlated with stress-induced ST elevation (≥0.5 mm) in right precordial leads, linking RV ischemia with proximal RCA involvement.^50^ Nakamura et al. further showed that in MI patients with interventricular septal (IVS) involvement, exercise RVEF was significantly lower (50±10%) compared to those without IVS involvement (56±9%), and abnormal RVCR was more frequent (68% vs. 36%).^51^ In a prospective study of patients with reperfused RV infarction, Karvounis et al. demonstrated that low-dose dobutamine stress echocardiography performed on day 5 post-MI identified stunned, viable myocardium, since the majority of hypokinetic segments improved wall motion with dobutamine, and these motility changes strongly predicted recovery of RV systolic and diastolic function at 3 months.^52^

In mitral stenosis (MS), Sade et al. used dobutamine stress echocardiography to show that MS patients had significantly lower RV isovolumic acceleration (IVA) at baseline (1.5±0.4 m/s²) and during dobutamine infusion (ΔIVA 1.5±0.4 m/s²) compared to controls (baseline: 2.4±0.5 m/s²; ΔIVA 3.2±0.7 m/s²). Similarly, RV S′ velocity was reduced in MS patients at baseline (8.1±1.4 cm/s) and during stress (10.3±1.9 cm/s), when compared with controls (baseline 10.3±1.7 cm/s; stress 15.5±2.6 cm/s). The +dP/dt/Pmax ratio was also significantly blunted in MS patients (baseline 6.2±1.4 1/s; stress 8.5±1.7 1/s) compared to controls (baseline 8.5±1.7 1/s; stress: 13.6±2.6 1/s). An IVA increase <3.4 m/s² independently predicted higher mortality and hospitalization rates. While mitral valve intervention improved RVCR, recovery was often incomplete compared to controls.^53^ In degenerative mitral regurgitation, Utsunomiya et al. demonstrated that patients with reduced exercise capacity (peak VO <70% predicted) had significantly lower TAPSE/sPAP slopes (0.10 mm/mmHg) compared to those with preserved capacity (0.46 mm/mmHg), indicating impaired RVCR. These patients also exhibited steeper mPAP/CO slopes (4.57 vs. 3.03 mmHg/L/min), reflecting reduced pulmonary vascular reserve.^54^ In asymptomatic patients with severe primary mitral regurgitation undergoing mitral valve surgery, Vitel et al. demonstrated that exercise TAPSE (ex-TAPSE) <26 mm was associated with significantly lower survival at 5 years. Additionally, patients with ex-TAPSE <26 mm experienced higher rates of major adverse cardiovascular events, including atrial fibrillation, stroke, cardiac-related hospitalization, and death.^55^

In patients with tricuspid regurgitation, Utsunomiya et al. developed a risk stratification system based on three-dimensional anatomical regurgitant orifice area (3D-AROA) and TAPSE/sPAP slope, which reflect the severity of regurgitation and RVCR, respectively. Patients were categorized into four groups based on 3D-AROA (<161 mm² or ≥161 mm²) and TAPSE/sPAP slope (>0.046 mm/mmHg or ≤0.046 mm/mmHg). Group IV (high risk: large 3D-AROA with impaired RVCR) had the worst outcomes, while Group I (low risk: smaller 3D-AROA with preserved RVCR) had the best survival.^56^

### 3.4 LVAD

Three studies including a total of 62 patients evaluated RVCR in LVAD patients with PV loops during exercise. A detailed summary of these studies is provided in **Table 4**. Ton et al. found that LVAD patients had the lowest augmentation in RVSWI (ΔRVSWI 1.5 L/min) compared to PAH (4.3 L/min) and controls (5.7 L/min), along with the greatest increase in RAP (ΔRAP 7 mmHg) and smallest CO augmentation (ΔCO 1.5 L/min) during exercise. RV afterload, assessed by Ea, increased significantly in LVAD patients (ΔEa 0.4 mmHg/mL) compared to PAH (0.1 mmHg/mL) and controls (0.1 mmHg/mL), correlating negatively with peak VO and ventilatory efficiency.^57^ Similarly, Tran et al. demonstrated limited RVCR in LVAD patients during submaximal exercise. Ees increased only from 0.37±0.14 mmHg/mL at rest to 0.52±0.32 mmHg/mL during exercise, while dP/dt_max_ rose modestly from 229±72 mmHg/sec to 246±75 mmHg/sec, with no significant augmentation at peak effort. These findings were associated with poor exercise tolerance and severe ventilatory inefficiency.^58^ Further analysis of RV systolic adaptation in LVAD patients demonstrated that those with lower ΔRVSWI (i.e., <2.08 g/m²) exhibited impaired RV contractile and diastolic reserve. Compared to patients with higher ΔRVSWI, they had significantly lower peak RV dP/dt_max_ (233 vs. 595 mmHg/sec) and less negative peak RV dP/dt_min_ (–269 vs. –465 mmHg/sec), indicating blunted contractile and lusitropic reserve. In addition, patients with lower ΔRVSWI demonstrated limited cardiac index augmentation (ΔCI 0.9 ± 0.4 L/min/m²) and reduced exercise tolerance. Importantly, ΔRVSWI correlated positively with both peak VO and 6-minute walk distance.^59^

**Table 4:** LVAD Patients. RV = Right Ventricle; RVCR = Right Ventricular Contractile Reserve; DRVSWI = Change in Right Ventricular Stroke Work Index; RVSWI = Right Ventricular Stroke Work Index; RHC = Right Heart Catheterization; CPET = Cardiopulmonary Exercise Testing; Δ (Delta) = Change; RV dP/dt = Rate of Rise of Right Ventricular Pressure; RAP = Right Atrial Pressure; CI = Cardiac Index; 6MWD = 6-Minute Walk Distance; HRQoL = Health-Related Quality of Life; PAC = Pulmonary Arterial Compliance; Ea = Effective Arterial Elastance (Pulmonary Arterial Load); CO = Cardiac Output; pVO[ = Peak Oxygen Uptake; VE/VCO[ Slope = Ventilatory Efficiency Slope; CF-LVAD = Continuous-Flow Left Ventricular Assist Device; Ees = End-Systolic Elastance (RV Contractility); Ees/Ea = RV-PA Coupling (Right Ventricle to Pulmonary Artery Coupling); PV Loop = Pressure-Volume Loop.

### 3.5 Other Conditions

RVCR was also studied in amyloidosis, iron deficiency, COPD, and cirrhosis. A detailed summary is provided in **Table 5**.

**Table 5:** Summary of Studies Assessing Right Ventricular Contractile Reserve in Various Conditions. RV = Right Ventricle; CR = Contractile Reserve; PH = Pulmonary Hypertension; HTDD = Heart Failure with Isolated Right Heart Disease; HFpEF = Heart Failure with Preserved Ejection Fraction; CTEPH = Chronic Thromboembolic Pulmonary Hypertension; RVESPAR = Right Ventricular End-Systolic Pressure-Area Ratio; TAPSE = Tricuspid Annular Plane Systolic Excursion; RVFWSRL = Right Ventricular Free Wall Strain Rate Longitudinal; PASP = Pulmonary Artery Systolic Pressure; RV-PA uncoupling = Right Ventricle to Pulmonary Artery Uncoupling; ID = Iron Deficiency; CPETecho = Cardiopulmonary Exercise Testing with Echocardiography; SPAP = Systolic Pulmonary Artery Pressure; CPET = Cardiopulmonary Exercise Testing; CMR = Cardiac Magnetic Resonance Imaging; RVEF = Right Ventricular Ejection Fraction; LVEF = Left Ventricular Ejection Fraction; SV = Stroke Volume; ECHO = Echocardiography; FAC = Fractional Area Change; S′ wave = Peak Systolic Velocity at the Tricuspid Annulus; ΔRVGLS = Change in Right Ventricular Global Longitudinal Strain; CMS = Chronic Mountain Sickness; HA = High Altitude; TDI = Tissue Doppler Imaging; ΔRV-S′ = Change in Right Ventricular Systolic Velocity (S′); ΔRV FAC = Change in Right Ventricular Fractional Area Change; ΔPASP = Change in Pulmonary Artery Systolic Pressure; VO[ = Oxygen Consumption; RVEDA = Right Ventricular End-Diastolic Area; TR grade = Tricuspid Regurgitation Grade; ΔGLS = Change in Global Longitudinal Strain; ΔGCS = Change in Global Circumferential Strain; GRS = Global Radial Strain; ΔEF = Change in Ejection Fraction; T1D = Type 1 Diabetes; RVLS = Right Ventricular Longitudinal Strain; ATTRwt-CA = Wild-Type Transthyretin Cardiac Amyloidosis; ΔRVSWI = Change in Right Ventricular Stroke Work Index; ΔPAWP/ΔCO = Change in Pulmonary Artery Wedge Pressure to Cardiac Output Ratio; NAC = National Amyloidosis Center Staging.

| Author (Year) | Aim of the Study | Study Population (n) | Testing Modality | Stress Modality | RV CR Parameters | Key Findings |
| --- | --- | --- | --- | --- | --- | --- |
| Takenaka et al. (2023) | Investigate the relationship between RV contractile reserve during exercise and exercise capacity in LVAD patients. | 24 LVAD patients (12 with lower ΔRVSWI, 12 with higher ΔRVSWI). | Invasive RHC | Supine cycling exercise. | ΔRVSWI<br>RV dP/dt<br>RAP<br>ΔCI | ΔRVSWI was positively correlated with 6MWD, peak $\dot{V}O_2$, and HRQoL. Patients with lower ΔRVSWI had reduced ΔCI and RV dP/dt. Resting RVSWI was not associated with exercise capacity or HRQoL. |
| Ton et al. (2021) | Assess RV reserve and adaptation to load during exercise in LVAD patients and compare with PAH and normal controls. | 12 LVAD patients, 12 PAH patients, and 12 normal controls. | Invasive RHC with CPET | Supine cycling exercise | RVSWI<br>PAC<br>Ea<br>ΔRAP<br>ΔCO | LVAD patients had the poorest RV reserve (lowest RVSWI and ΔCO, highest ΔRAP). Exercise-induced RV afterload (Ea) correlated strongly with poor exercise capacity (low $p\dot{V}O_2$) and ventilatory inefficiency (high $VE/VCO_2$ slope), highlighting impaired RV reserve compared to PAH and controls. |
| Tran et al. (2021) | Characterize RV function during exercise and its impact on functional capacity in CF-LVAD patients. | 26 CF-LVAD patients (13 conductance catheter, 13 Swan-Ganz) | Invasive RHC with conductance or Swan-Ganz catheters | Upright cycle ergometry | Ees<br>Ea<br>Ees/Ea ratio<br>ΔCO<br>Δdp/dt<br>RV PV loop area | CF-LVAD patients demonstrated limited RV reserve with modest increases in contractility (Ees, Δdp/dt) during sub-maximal exercise but no significant changes at peak exercise. Persistent RV dysfunction with elevated filling pressures impaired functional capacity and exercise tolerance. |
| Claeys et al. (2022) | Investigate RV CR and PV load in HTDD, HFpEF, and CTEPH patients during exercise. | 53 subjects: 10 controls, 13 HTDD, 15 HFpEF, 15 CTEPH. | Exercise stress echocardiography | Semi-supine cycling exercise | RVESPAR<br>TAPSE<br>RVFWSRL<br>PASP | RV CR, measured via RVESPAR, was preserved in controls and HTDD but borderline in HFpEF and significantly reduced in CTEPH. HFpEF and CTEPH exhibited RV-PA uncoupling, with increased PASP and reduced TAPSE. HTDD display intermediate findings, with reduced RV systolic reserve compared to controls. |
| Martens et al. (2021) | Assess the impact of iron deficiency on RV contractile reserve in patients with unexplained dyspnea. | 272 subjects: 209 without ID and 63 with ID. | Cardiopulmonary exercise testing and echocardiography (CPETecho) | Upright cycling exercise | TAPSE/SPAP<br>RVESPAR | Patients with ID exhibited significantly impaired RV CR, with lower RVESPAR and a higher proportion showing RV-PA uncoupling during exercise (TAPSE/SPAP ratio < 0.36). These findings suggest that ID is associated with reduced biventricular reserve and exercise capacity. |
| Patterson et al. (2020) | Evaluate dyspnea of unknown etiology in HIV patients | 34 HIV patients with dyspnea on exertion | CPET and CMR | Treadmill exercise | RVEF<br>LVEF<br>SV changes | Preserved RV and LV contractile reserves (mean absolute RV CR: 9.0%). No exercise-induced PAH or significant scar burden. Impaired ventilation suggests a non-cardiopulmonary etiology for dyspnea, possibly subclinical myopathy. |
| Pergola et al. (2021) | Assess the feasibility and utility of RV stress echocardiography in adult patients. | 81 patients referred for stress ECHO | Stress ECHO | Exercise or dobutamine<br>Specific infusion doses were not reported | TAPSE<br>FAC<br>S' wave<br>sPAP<br>ΔRVGLS | RV contractile reserve assessed by ΔRVGLS increased by 39.8% with stress. Stress echocardiography was highly feasible, with reliable assessment of TAPSE, S-wave, and sPAP. ΔRVGLS was valuable for RVCR assessment in patients with good acoustic windows. |
| Pratali et al. (2013) | To evaluate RV and LV function and pulmonary circulation in chronic mountain sickness (CMS) patients compared with healthy high-altitude (HA) dwellers. | 86 participants (46 CMS, 40 HA) | Doppler echocardiography and TDI | Semi-supine bicycle exercise | RV-ESPAR<br>ΔRV-S'<br>ΔRV FAC | The study concluded that RV contractile reserve was preserved in both CMS patients and HA dwellers, as demonstrated by a significant increase in RV-ESPAR during exercise. While CMS patients exhibited lower resting RV function, including reduced FAC and RV-S', the exercise-induced improvements in contractility were comparable between the groups. |
| Rodríguez et al. (2017) | Assess RV CR in severe COPD patients during exercise using PASP as a marker. | 81 patients with severe COPD (66 male, 15 female). | CPET with echocardiography | Supine cycling exercise | ΔPASP<br>VO<sub>2</sub> | ΔPASP ≥ 30 mmHg was associated with higher peak VO<sub>2</sub> and better survival. ΔPASP < 30 mmHg identified patients with reduced exercise capacity and higher mortality risk. Exercise-induced PASP was identified as a prognostic factor for survival in COPD. |
| Rozenbaum et al. (2021) | Investigate determinants of exercise limitation in COPD patients using combined CPET and stress echocardiography. | 40 participants: 20 COPD patients, 20 controls. | Combined CPET and stress ECHO | Semi-supine cycling exercise | RV S'<br>RVEDA<br>RVFAC<br>TR grade | COPD patients exhibited abnormal RV systolic reserve, blunted SV increase, septal flattening, reduced LV compliance, and increased LV filling pressures during exercise. Limited SV reserve and chronotropic incompetence were major contributors to effort intolerance. |
| Sampaio et al. (2015) | Investigate systolic response to dobutamine in patients with cirrhotic cardiomyopathy. | 36 cirrhosis patients, 8 healthy controls. | Dobutamine stress cardiovascular MRI<br>Dobutamine was infused at 10 μg/kg/min for 3 minutes, then increased to 20 μg/kg/min for 3 minutes. | Pharmacologic dobutamine stress (10-20 μg/kg/min) | ΔGLS<br>ΔGCS<br>GRS<br>ΔEF | Cirrhotic patients demonstrated blunted RV CR with reduced ΔGLS, ΔGCS, and ΔEF: 10 μg/kg/min compared to controls. Improvements in strain metrics normalized to higher doses. |
| Zimmermann et al. (2023) | Investigate dynamic cardiac remodeling of the right and left heart in asymptomatic individuals with type 1 diabetes. | 8 asymptomatic T1D participants (5 males, 3 females; aged 23–65 years). | 2D echocardiography with speckle-tracking analysis. | Peak exercise on a bicycle. | ΔRV strain | In asymptomatic T1D patients, ΔRV strain showed no significant change after exercise, indicating preserved RV contractile function at rest and post-exercise. |
| Agudo et al. (2022) | Assess short-term prognosis in ATTRwt-CA patients using RV CR. | 11 ATTRwt-CA patients: Group 1 (n=6) with CV events; Group 2 (n=5) event-free. | Exercise ECHO | Upright bicycle exercise | RVGLS<br>Free-wall RVLS<br>TAPSE<br>RV S' | Group 1 showed no RV CR and worse RV function (lower RVGLS, RVLS, TAPSE, and RV S') compared to Group 2. Group 2 had preserved RV CR, better RV function, and higher free-wall RVLS value (CR by free-wall RVLS), indicating strong prognostic value for event-free survival. |
| Ladefoged et al. (2024) | To investigate the association between exercise hemodynamics, mitochondrial oxidative capacity, and disease stage in wild-type transthyretin amyloid cardiomyopathy (ATTRwt). | 47 ATTRwt patients (NAC I-L: 14, NAC I-H: 20, NAC II-III: 13) | Right heart catheterization | Cardiopulmonary exercise testing | ΔRVSWI<br>ΔPAWP/ΔCO | RV contractile reserve (ΔRVSWI) was reduced at peak exercise in patients with advanced ATTRwt stages. Patients with NAC II-III had significantly lower ΔRVSWI compared to NAC I-L. RV function was impaired during exercise, showing abnormal restrictive physiology even in early-stage |
| | | | | | | ATTRwt. A progressive impairment of the pressure-flow relationship (ΔPAWP/ΔCO) was noted across disease stages, with the most pronounced abnormality in NAC II-I patients. |

#### Amyloid Cardiomyopathy

Agudo et al. studied 11 patients with biopsy-proven wild-type transthyretin amyloidosis (ATTRwt) using upright bicycle ergometry.^60^ Their findings demonstrated RVGLS and RVFWS showed stronger associations with cardiovascular death and hospitalizations for heart failure compared to LV contractile reserve. Specifically, patients without events had more negative RVGLS (–17.3±1.1% vs. –9.5±0.9%) and RVFWS (–18.3±1.0% vs. –8.8±1.0%) during exercise, with RVFWS emerging as the strongest predictor of event-free survival.

Ladefoged et al. investigated 47 patients with biopsy-confirmed ATTRwt, stratified by National Amyloidosis Centre (NAC) staging (I-L, I-H, II/III), using exercise RHC. Abnormal biventricular responses to exercise were observed even in early-stage patients (NAC I-L), who had normal resting filling pressures but developed elevated RAP, PAWP, and mPAP during exercise. Despite these pressure increases, CO augmentation was blunted, particularly in advanced stages (NAC I-H, II/III). Peak CO in NAC I-L was 9.4 L/min, compared to 7.0 L/min in NAC I-H and 6.3 L/min in NAC II/III. Stroke volume did not significantly increase across groups. RVSWI responses were also diminished with advancing disease (peak RVSWI: 15±6 g*m/m² in NAC I-L vs. 10±5 g*m/m² in NAC I-H and 9±5 g*m/m² in NAC II/III). Additionally, pressure-flow relationships worsened with disease progression, as seen with elevated PAWP/CO (rest: 2.7 to 4.4 WU across stages) and PAWP/Watt ratios. These findings suggest that RVCR impairment begins early in ATTRwt and progressively worsens, contributing to reduced exercise capacity.^61^

#### Chronic Obstructive Pulmonary Disease

Berger et al. studied sixteen COPD patients using radionuclide angiocardiography during upright bicycle exercise. Twelve patients out of sixteen showed blunted RVEF responses.^62^ Furthermore, COPD severity (assessed with FEV_1_ and resting PaO_2_), correlated with the degree of RVCR impairment, although no differences in workload achieved were observed between patients with normal versus abnormal RV reserve.^62^ Rodríguez et al. evaluated 81 patients with severe COPD using CPET with simultaneous Doppler echocardiography. They found that patients with an exercise-induced SPAP increase ≤30 mmHg (n=16) had significantly shorter survival over 3 years compared to those with increases >30 mmHg (n=65).^63^

More recently, Rozenbaum et al. studied twenty patients with mild-moderate COPD using combined CPET and semi-supine exercise echocardiography.^64^ Compared to controls, COPD patients demonstrated blunted increases in RV S′ (from 5.6±1.9 to 6.5±2.5 cm/sec in COPD vs. 7.1±1.8 to 9.3±3.7 cm/sec in controls) and RVFAC (from 39.7±11% to 42.2±11% vs. 47.3±9% to 66.5±15%, respectively). COPD patients also showed no significant increase in SV during exercise (ΔSV –0.5±15.8 mL) compared to substantial augmentation in controls (ΔSV 16.9±19.6 mL), contributing to reduced CO during exercise and exercise intolerance. These findings are consistent with Berger et al., who demonstrated no increase in RVEF during upright exercise, similar to CAD patients, despite preserved resting RVEF.^62^

#### Iron Deficiency

Martens et al. evaluated 272 patients using exercise CMR and echocardiography, comparing those with iron deficiency (n = 63) to those without (n = 209).^65^ Iron deficiency was associated with reduced RV and LV contractile reserve as well as RV-PA uncoupling during exercise, as shown by lower TAPSE, TAPSE/sPAP, and RVESPAR. These impairments occurred despite similar resting and exercise filling pressures between the two groups, suggesting a direct myocardial effect of iron deficiency. Even after adjusting for LV contractile reserve, iron deficiency remained independently associated with blunted RVCR.^65^

#### Cirrhotic Cardiomyopathy

Sampaio et al. compared 36 patients with cirrhosis to eight controls using dobutamine stress CMR.^66^ Despite similar resting parameters, cirrhotic patients exhibited blunted increases in SV, CO, and strain (RVGLS and RVGCS) during stress. Specifically with strain analysis, significant differences were observed in RVGLS (-1.4 vs. -5.9) and RVGCS (-4.0 vs. -8.3).^66^ *Type 1 Diabetes*

Diabetes mellitus independently increases the long-term risk of heart failure, even in the absence of CAD or hypertension. However, Zimmermann et al. found that in a small cohort of asymptomatic adults with type 1 diabetes, RV function, assessed by speckle-tracking strain at rest and post-exercise, was preserved.^67^

#### HIV-Associated Cardiomyopathy

HIV has been shown to be an independent risk factor for heart failure, though little evidence exists regarding its specific effects on RV function.^68^ Patterson et al. examined 34 HIV patients with unexplained dyspnea using CPET and exercise CMR.^69^ The mean absolute RVCR was preserved during exercise (ΔRVEF and ΔRVSV were 9.0%±11.2% and 7.2±25.3 ml, respectively), with no evidence of exercise-induced PH, suggesting that RV dysfunction was not a major contributor to dyspnea in this population.^69^

## Discussion

This scoping review synthesizes current approaches to the assessment of RVCR across a broad spectrum of populations, including healthy individuals and athletes, patients with PH, structural and functional heart disease, LVAD recipients, and other conditions. Across modalities and medical conditions, impaired RVCR was consistently associated with adverse outcomes, yet variability in definitions, protocols, and type of stress modalities limits its clinical adoption.

Echocardiography was the most studied modality for assessing RVCR. Stress-induced changes in TAPSE, RVGLS, and RVFWLS were frequently evaluated. Blunted responses, typically a TAPSE increase <3–5 mm, RVGLS improvement <4–5%, or RVFWLS change <5%, were linked to reduced exercise capacity, worse survival, and impaired RV–PA coupling, even when resting RV function appeared normal. Strain parameters, particularly RVGLS and RVFWLS, showed moderately strong correlation with CMR-derived RVEF, improving the diagnostic accuracy of echocardiography, adding an 8% net reclassification improvement beyond conventional measures like TAPSE or S′.^70^ Strain imaging has been shown to be sensitive for detecting subclinical right ventricular dysfunction across diverse populations, including patients with pulmonary hypertension and advanced heart failure. Studies demonstrate that strain-based indices, particularly RVGLS, are less affected by confounding factors like abnormal longitudinal rotation.^71,72^ Importantly, RVGLS offers incremental prognostic value beyond clinical and hemodynamic markers and meaningfully improves risk stratification. This is especially relevant in predicting outcomes such as right ventricular failure following LVAD implantation.^73^ Additionally, elevated peak sPAP (>50–60 mmHg) and a reduced TAPSE/sPAP ratio, especially <0.36 mm/mmHg at peak exercise, were also strongly associated with impaired RV–PA coupling and worse outcomes in pulmonary hypertension and heart failure.

RHC and PV loop analysis provide invasive hemodynamic insights into RVCR. RHC was more commonly used and often integrated with exercise or pharmacologic stress (e.g., dobutamine), and in some cases combined with echocardiography. Parameters such as Ea, ΔRAP, ΔCO, ΔmPAP/ΔCO slope, and TAPSE/sPAP were frequently reported. Impaired RVCR in general was associated with higher Ea (>0.5–0.8 mmHg/mL), greater ΔRAP (>5 mmHg), and blunted ΔCO (<2.0 L/min) during exercise or pharmacologic stress, indicating inadequate contractile adaptation. A steeper ΔmPAP/ΔCO slope (>3 mmHg/L/min) reflected reduced pulmonary vascular reserve, while a lower TAPSE/sPAP ratio, particularly <0.36 mm/mmHg at peak exercise, was linked to RV–PA uncoupling and adverse outcomes. These abnormalities were particularly evident in patients with PH, SSc, HF, and LVAD recipients. While RHC enables detailed pressure–flow assessment, it lacks volumetric data and cannot directly measure Ees or Ees/Ea. PV loop analysis, though less frequently performed, remains the gold standard and demonstrated robust increases in RV stroke work index (RVSWI) and Ees in healthy individuals, whereas patients with PH, HF, and LVADs exhibited blunted Ees augmentation and reduced Ees/Ea (typically <1.0), indicating impaired RVCR and RV–PA uncoupling.

CMR, although used in fewer studies, demonstrated strong concordance with invasive findings. A decline in RVEF during submaximal exercise, particularly a failure to increase by ≥20% from baseline or a drop below 38%, reflected impaired RV adaptation to rising afterload. CMR-derived strain parameters, including RVGLS and RVFWLS, were more sensitive than RVEF in detecting subclinical dysfunction. Blunted strain augmentation, defined as an increase in RVGLS or RVFWLS of <4–5% (i.e., more negative) from baseline, was observed in patients with PAH and other conditions such as those with LVADs. RV strain measures correlated inversely with Ees and Ees/Ea, while circumferential strain aligned with Ea, reinforcing the physiologic relevance of CMR-based metrics.^74^ Additionally, CMR-derived TAPSE/sPAP and mPAP/CO slopes showed close agreement with invasive PV loop measurements, with impaired RV–PA coupling typically indicated by TAPSE/sPAP <0.36 mm/mmHg and a ΔmPAP/ΔCO slope >3 mmHg/L/min.

The choice of stress modality, exercise (supine or upright) versus dobutamine, was addressed in relatively few studies. Exercise most closely replicates physiologic loading conditions, typically leading to pulmonary vasodilation and vascular recruitment that reduce afterload. However, in patients with pulmonary vascular disease, exertion may still unmask abnormal increases in afterload due to impaired vascular reserve. Dobutamine, by contrast, more selectively isolates inotropic reserve, but its hemodynamic effects depend on dose: lower doses (5–10 μg/kg/min) primarily stimulate β1-receptors to increase contractility, while higher doses (≥20 μg/kg/min) can activate α-receptors and induce pulmonary vasoconstriction, potentially confounding RV afterload assessment. Despite these differences, few studies directly compared these stressors or standardized protocols across populations, highlighting an important area for future research.

This review is limited by substantial heterogeneity in study design, patient populations, stress modalities, and RVCR definitions. Most studies were observational, single-center, and small, introducing the potential for selection and reporting bias. Sample sizes varied, and outcome measures were inconsistently reported. Technical variability, particularly in strain analysis, further complicates reproducibility. Importantly, no consensus exists for defining abnormal RVCR, and few studies offered direct comparisons between imaging modalities or stress protocols. Nevertheless, despite the absence of universally accepted thresholds, **Table 6** summarizes proposed cutoff values for preserved RVCR parameters during stress based on the available literature. Finally, no prospective studies evaluated RVCR-guided management strategies, leaving its role in clinical decision-making unclear.

**Table 6.**
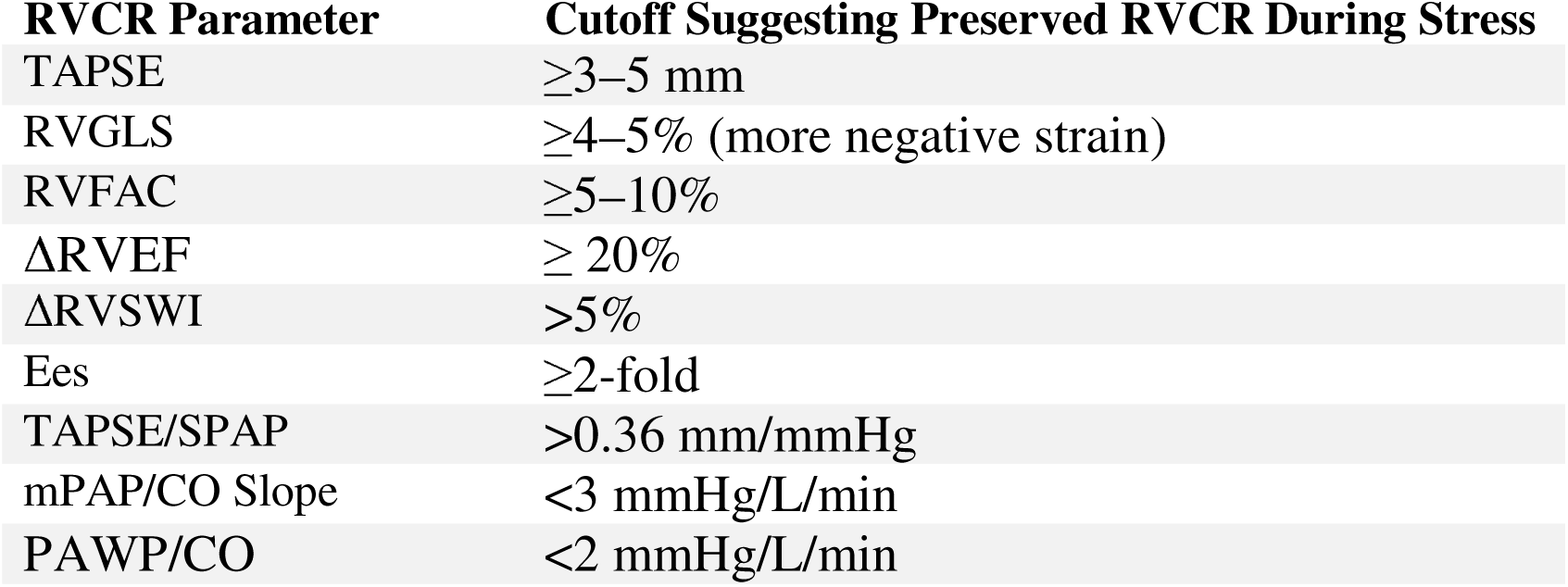
Proposed cutoff values suggesting preserved RVCR during stress.

| RVCR Parameter | Cutoff Suggesting Preserved RVCR During Stress |
| --- | --- |
| TAPSE | $\geq 3\text{--}5$ mm |
| RVGLS | $\geq 4\text{--}5\%$ (more negative strain) |
| RVFAC | $\geq 5\text{--}10\%$ |
| ΔRVEF | $\geq 20\%$ |
| ΔRVSWI | $> 5\%$ |
| Ees | $\geq 2$-fold |
| TAPSE/SPAP | $> 0.36$ mm/mmHg |
| mPAP/CO Slope | $< 3$ mmHg/L/min |
| PAWP/CO | $< 2$ mmHg/L/min |

Based on our findings, several key areas warrant further investigation. First, standardized definitions and thresholds for abnormal RVCR across imaging modalities and stress protocols are needed. Second, prospective, multicenter studies are required to validate the prognostic utility of RVCR-guided clinical strategies, determining whether RVCR assessment can inform therapeutic interventions and timing to improve patient outcomes. Comparative studies evaluating invasive versus non-invasive methods could clarify modality-specific advantages and clinical applicability. Lastly, novel interventions or pharmacologic strategies designed specifically to enhance RVCR deserve investigation, particularly in populations such as PH, HF, and LVAD recipients, where impaired RVCR profoundly influences morbidity and mortality. Addressing these gaps will likely enable more precise patient risk stratification and improve management and hence outcomes.

## Conclusion

Right ventricular contractile reserve is a clinically relevant marker of right heart adaptability across a broad spectrum of cardiopulmonary diseases, providing prognostic information beyond what is offered by resting values. While multiple modalities have demonstrated its utility, the lack of standardized protocols and validated thresholds limits its integration into clinical practice. Prospective, multicenter studies are needed to establish best practices, define abnormal RVCR, and determine whether RVCR-guided strategies can improve patient outcomes.

## Supporting information

Online supplement - search strategy

## Data Availability

All data produced are available in the original articles included in this review.

## Abbreviations Used in the Manuscript

Abbreviation: Full Term
ARDS: acute fespiratory distress syndrome
CAD: coronary artery disease
CMR: cardiac magnetic resonance imaging
COPD: chronic obstructive pulmonary disease
CO: cardiac output
CPET: cardiopulmonary exercise testing
CTED: chronic thromboembolicdisease
CTEPH: chronic thromboembolic pulmonary hypertension
Ea: arterial elastance
Ees: end-systolic elastance
Ees/Ea: Right ventricle–pulmonary artery coupling ratio
ePVH: exercise pulmonary venous hypertension
FAC: fractional area change
FWLS: free wall longitudinal strain
HF: heart failure
HFpEF: heart failure with preserved ejection fraction
HFrEF: heart failure with reduced ejection fraction
HCM: hypertrophic cardiomyopathy
IDC: idiopathic dilated cardiomyopathy
IPF: idiopathic pulmonary fibrosis
IVS: interventricular septum
LV: left ventricle
LVAD: left ventricular assist device
LVEF: left ventricular ejection fraction
MI: myocardial infarction
MS: mitral stenosis
mPAP: mean pulmonary artery pressure
mPAP/CO Slope: pressure–flow relationship during exercise
PAH: pulmonary arterial hypertension
PAP: pulmonary artery pressure
PAPS: pulmonary artery pressure systolic
PASP: pulmonary artery systolic pressure
PH: pulmonary hypertension
PTAS: pulmonary transit of agitated saline
PV: pressure–volume
PVR: pulmonary vascular resistance
RHC: right heart catheterization
RV: right ventricle
RVCR: right ventricular contractile reserve
RVEDA: right ventricular end-diastolic area
RVESPAR: right ventricular end-systolic pressure–area ratio
RVEF: right ventricular ejection fraction
RVFAC: right ventricular fractional area change
RVFWLS: right ventricular free wall longitudinal strain
RVGLS: right ventricular global longitudinal strain
RVLSS: right ventricular longitudinal systolic strain
RVSP: right ventricular systolic pressure
RVSW: right ventricular stroke work
RVSWI: right ventricular stroke work index
S′: tricuspid annular systolic velocity
SSc: systemic sclerosis
SV: stroke volume
TAPSE: tricuspid annular plane systolic excursion
TP/ET: time-to-peak to ejection time ratio
TR: tricuspid regurgitation
VO: oxygen consumption
WU: wood units

## References

1. Haddad F, Doyle R, Murphy DJ, Hunt SA. Right Ventricular Function in Cardiovascular Disease, Part II: Pathophysiology, Clinical Importance, and Management of Right Ventricular Failure. Circulation. 2008;117(13):1717–1731. doi:10.1161/CIRCULATIONAHA.107.653584

2. Voelkel NF, Quaife RA, Leinwand LA, et al. Right Ventricular Function and Failure: Report of a National Heart, Lung, and Blood Institute Working Group on Cellular and Molecular Mechanisms of Right Heart Failure. Circulation. 2006;114(17):1883–1891. doi:10.1161/CIRCULATIONAHA.106.632208

3. Gorter TM, Van Melle JP, Rienstra M, et al. Right Heart Dysfunction in Heart Failure With Preserved Ejection Fraction: The Impact of Atrial Fibrillation. J Card Fail. 2018;24(3):177–185. doi:10.1016/j.cardfail.2017.11.005

4. Hassoun PM. Pulmonary Arterial Hypertension. Taichman DB, ed. N Engl J Med. 2021;385(25):2361–2376. doi:10.1056/NEJMra2000348

5. Konstam MA, Kiernan MS, Bernstein D, et al. Evaluation and Management of Right-Sided Heart Failure: A Scientific Statement From the American Heart Association. Circulation. 2018;137(20). doi:10.1161/CIR.0000000000000560

6. Sato R, Dugar S, Cheungpasitporn W, et al. The impact of right ventricular injury on the mortality in patients with acute respiratory distress syndrome: a systematic review and meta-analysis. Crit Care. 2021;25(1):172. doi:10.1186/s13054-021-03591-9

7. Sanz J, Sánchez-Quintana D, Bossone E, Bogaard HJ, Naeije R. Anatomy, Function, and Dysfunction of the Right Ventricle. J Am Coll Cardiol. 2019;73(12):1463–1482. doi:10.1016/j.jacc.2018.12.076

8. Cornwell WK, Tran T, Cerbin L, et al. New insights into resting and exertional right ventricular performance in the healthy heart through real-time pressure-volume analysis. J Physiol. 2020;598(13):2575–2587. doi:10.1113/JP279759

9. Houston BA, Brittain EL, Tedford RJ. Right Ventricular Failure. Taichman DB, ed. N Engl J Med. 2023;388(12):1111–1125. doi:10.1056/NEJMra2207410

10. D’Alto M, Pavelescu A, Argiento P, et al. Echocardiographic assessment of right ventricular contractile reserve in healthy subjects. Echocardiogr Mt Kisco N. 2017;34(1):61–68. doi:10.1111/echo.13396

11. La Gerche A, Burns AT, D’Hooge J, MacIsaac AI, Heidbuchel H, Prior DL. Exercise Strain Rate Imaging Demonstrates Normal Right Ventricular Contractile Reserve and Clarifies Ambiguous Resting Measures in Endurance Athletes. J Am Soc Echocardiogr. doi:10.1016/j.echo.2011.11.023

12. Sanz-de la Garza M, Vaquer-Segui A, Duran K, et al. Pulmonary transit of contrast during exercise is related to improved cardio-pulmonary performance in highly trained endurance athletes. Eur J Prev Cardiol. 2020;27(14):1504–1514. doi:10.1177/2047487319891779

13. Blumberg FC, Arzt M, Lange T, Schroll S, Pfeifer M, Wensel R. Impact of right ventricular reserve on exercise capacity and survival in patients with pulmonary hypertension. Eur J Heart Fail. 2013;15(7):771–775. doi:10.1093/eurjhf/hft044

14. Grunig E, Tiede H, Enyimayew EO, et al. Assessment and prognostic relevance of right ventricular contractile reserve in patients with severe pulmonary hypertension. Circulation. 2013;128(18):2005–2015. doi:10.1161/CIRCULATIONAHA.113.001573

15. Sano H, Tanaka H, Motoji Y, et al. Echocardiography during preload stress for evaluation of right ventricular contractile reserve and exercise capacity in pulmonary hypertension. Echocardiogr Mt Kisco N. 2018;35(12):1997–2004. doi:10.1111/echo.14161

16. Spruijt OA, de Man FS, Groepenhoff H, et al. The effects of exercise on right ventricular contractility and right ventricular-arterial coupling in pulmonary hypertension. Am J Respir Crit Care Med. 2015;191(9):1050–1057. doi:10.1164/rccm.201412-2271OC

17. Almeida AR, Loureiro MJ, Cotrim C, Lopes L, Repolho D, Pereira H. Echocardiographic assessment of right ventricular contractile reserve in patients with pulmonary hypertension. Rev Port Cardiol. 2014;33(3):155–163. doi:10.1016/j.repc.2013.09.015

18. Guo DC, Li YD, Yang YH, et al. Influence of impaired right ventricular contractile reserve on exercise capacity in patients with precapillary pulmonary hypertension: A study with exercise stress echocardiography. Echocardiogr Mt Kisco N. 2019;36(4):671–677. doi:10.1111/echo.14283

19. Sharma T, Lau EMT, Choudhary P, et al. Dobutamine stress for evaluation of right ventricular reserve in pulmonary arterial hypertension. Eur Respir J. 2015;45(3):700–708. doi:10.1183/09031936.00089914

20. Jaijee S, Quinlan M, Tokarczuk P, et al. Exercise cardiac MRI unmasks right ventricular dysfunction in acute hypoxia and chronic pulmonary arterial hypertension. Am J Physiol Heart Circ Physiol. 2018;315(4):H950–H957. doi:10.1152/ajpheart.00146.2018

21. Singh I, Rahaghi FN, Naeije R, et al. Dynamic right ventricular-pulmonary arterial uncoupling during maximum incremental exercise in exercise pulmonary hypertension and pulmonary arterial hypertension. Pulm Circ. 2019;9(3):2045894019862435. doi:10.1177/2045894019862435

22. Ireland CG, Damico RL, Kolb TM, et al. Exercise right ventricular ejection fraction predicts right ventricular contractile reserve. J Heart Lung Transplant Off Publ Int Soc Heart Transplant. 2021;40(6):504–512. doi:10.1016/j.healun.2021.02.005

23. Chia EM, Lau EMT, Xuan W, Celermajer DS, Thomas L. Exercise testing can unmask right ventricular dysfunction in systemic sclerosis patients with normal resting pulmonary artery pressure. Int J Cardiol. 2016;204:179–186. doi:10.1016/j.ijcard.2015.11.186

24. Mukherjee M, Mercurio V, Hsu S, et al. Assessment of right ventricular reserve utilizing exercise provocation in systemic sclerosis. Int J Cardiovasc Imaging. 2021;37(7):2137–2147. doi:10.1007/s10554-021-02237-9

25. Rallidis LS, Papangelopoulou K, Anthi A, et al. The Role of Exercise Doppler Echocardiography to Unmask Pulmonary Arterial Hypertension in Selected Patients with Systemic Sclerosis and Equivocal Baseline Echocardiographic Values for Pulmonary Hypertension. Diagn Basel Switz. 2021;11(7). doi:10.3390/diagnostics11071200

26. Hsu S, Houston BA, Tampakakis E, et al. Right Ventricular Functional Reserve in Pulmonary Arterial Hypertension. Circulation. 2016;133(24):2413–2422. doi:10.1161/CIRCULATIONAHA.116.022082

27. Xanthouli P, Miazgowski J, Benjamin N, et al. Prognostic meaning of right ventricular function and output reserve in patients with systemic sclerosis. Arthritis Res Ther. 2022;24(1):173. doi:10.1186/s13075-022-02863-1

28. Claeys M, Claessen G, La Gerche A, et al. Impaired Cardiac Reserve and Abnormal Vascular Load Limit Exercise Capacity in Chronic Thromboembolic Disease. JACC Cardiovasc Imaging. 2019;12(8 Pt 1):1444–1456. doi:10.1016/j.jcmg.2018.07.021

29. Claessen G, La Gerche A, Voigt JU, et al. Accuracy of Echocardiography to Evaluate Pulmonary Vascular and RV Function During Exercise. JACC Cardiovasc Imaging. 2016;9(5):532–543. doi:10.1016/j.jcmg.2015.06.018

30. Claessen G, La Gerche A, Dymarkowski S, Claus P, Delcroix M, Heidbuchel H. Pulmonary vascular and right ventricular reserve in patients with normalized resting hemodynamics after pulmonary endarterectomy. J Am Heart Assoc. 2015;4(3):e001602. doi:10.1161/JAHA.114.001602

31. Braams NJ, Kianzad A, Meijboom LJ, et al. Right Ventricular Function During Exercise After Pulmonary Endarterectomy for Chronic Thromboembolic Pulmonary Hypertension. J Am Heart Assoc. 2023;12(4):e027638. doi:10.1161/JAHA.122.027638

32. D’Andrea A, Stanziola AA, Saggar R, et al. Right Ventricular Functional Reserve in Early-Stage Idiopathic Pulmonary Fibrosis: An Exercise Two-Dimensional Speckle Tracking Doppler Echocardiography Study. Chest. 2019;155(2):297–306. doi:10.1016/j.chest.2018.11.015

33. McCabe C, Oliveira RKF, Rahaghi F, et al. Right ventriculo-arterial uncoupling and impaired contractile reserve in obese patients with unexplained exercise intolerance. Eur J Appl Physiol. 2018;118(7):1415–1426. doi:10.1007/s00421-018-3873-4

34. Bandera F, Generati G, Pellegrino M, et al. Role of right ventricle and dynamic pulmonary hypertension on determining DELTAVO2/DELTAWork Rate flattening: insights from cardiopulmonary exercise test combined with exercise echocardiography. Circ Heart Fail. 2014;7(5):782–790. doi:10.1161/CIRCHEARTFAILURE.113.001061

35. Lin ACW, Seale H, Hamilton-Craig C, Morris NR, Strugnell W. Quantification of biventricular strain and assessment of ventriculo-ventricular interaction in pulmonary arterial hypertension using exercise cardiac magnetic resonance imaging and myocardial feature tracking. J Magn Reson Imaging JMRI. 2019;49(5):1427–1436. doi:10.1002/jmri.26517

36. Romeih S, Blom NA, Van der Plas MN, et al. Impaired cardiac reserve in asymptomatic patients with moderate pulmonary restenosis late after relief of severe pulmonary stenosis: evidence for diastolic dysfunction. Int J Cardiol. 2013;167(6):2836–2840. doi:10.1016/j.ijcard.2012.07.029

37. Borlaug BA, Kane GC, Melenovsky V, Olson TP. Abnormal right ventricular-pulmonary artery coupling with exercise in heart failure with preserved ejection fraction. Eur Heart J. 2016;37(43):3293–3302. doi:10.1093/eurheartj/ehw241

38. Singh I, Rahaghi FN, Naeije R, Oliveira RKF, Systrom DM, Waxman AB. Right Ventricular-Arterial Uncoupling During Exercise in Heart Failure With Preserved Ejection Fraction: Role of Pulmonary Vascular Dysfunction. Chest. 2019;156(5):933–943. doi:10.1016/j.chest.2019.04.109

39. Rommel KP, von Roeder M, Oberueck C, et al. Load-Independent Systolic and Diastolic Right Ventricular Function in Heart Failure With Preserved Ejection Fraction as Assessed by Resting and Handgrip Exercise Pressure-Volume Loops. Circ Heart Fail. 2018;11(2):e004121. doi:10.1161/CIRCHEARTFAILURE.117.004121

40. Guazzi M, Villani S, Generati G, et al. Right Ventricular Contractile Reserve and Pulmonary Circulation Uncoupling During Exercise Challenge in Heart Failure: Pathophysiology and Clinical Phenotypes. JACC Heart Fail. 2016;4(8):625–635. doi:10.1016/j.jchf.2016.03.007

41. Gorcsan IJ, Murali S, Counihan PJ, Mandarino WA, Kormos RL. Right ventricular performance and contractile reserve in patients with severe heart failure: Assessment by pressure-area relations and association with outcome. Circulation. 1996;94(12):3190–3197. doi:10.1161/01.CIR.94.12.3190

42. Kinoshita M, Saito M, Inoue K, et al. Role of the right ventricular contractile reserve during low-load exercise in predicting heart failure readmission. J Cardiol. 2023;82(1):43–50. doi:10.1016/j.jjcc.2023.03.001

43. Kinoshita M, Inoue K, Higashi H, et al. Impact of right ventricular contractile reserve during low-load exercise on exercise intolerance in heart failure. ESC Heart Fail. 2020;7(6):3810–3820. doi:10.1002/ehf2.12968

44. D’Andrea A, Limongelli G, Baldini L, et al. Exercise speckle-tracking strain imaging demonstrates impaired right ventricular contractile reserve in hypertrophic cardiomyopathy. Int J Cardiol. 2017;227:209–216. doi:10.1016/j.ijcard.2016.11.150

45. Wu XP, Li YD, Wang YD, et al. Impaired Right Ventricular Mechanics at Rest and During Exercise Are Associated With Exercise Capacity in Patients With Hypertrophic Cardiomyopathy. J Am Heart Assoc. 2019;8(5):e011269. doi:10.1161/JAHA.118.011269

46. Plehn G, Vormbrock J, Perings S, et al. Comparison of right ventricular functional response to exercise in hypertrophic versus idiopathic dilated cardiomyopathy. Am J Cardiol. 2010;105(1):116–121. doi:10.1016/j.amjcard.2009.08.662

47. Otasevic P, Popovic Z, Pratali L, Vlahovic A, Vasiljevic JD, Neskovic AN. Right vs. left ventricular contractile reserve in one-year prognosis of patients with idiopathic dilated cardiomyopathy: assessment by dobutamine stress echocardiography. Eur J Echocardiogr J Work Group Echocardiogr Eur Soc Cardiol. 2005;6(6):429–434. doi:10.1016/j.euje.2005.01.006

48. Matsumoto K, Tanaka H, Onishi A, et al. Bi-ventricular contractile reserve offers an incremental prognostic value for patients with dilated cardiomyopathy. Eur Heart J Cardiovasc Imaging. 2015;16(11):1213–1223. doi:10.1093/ehjci/jev069

49. Berger HJ, Johnstone DE, Sands JM, Gottschalk A, Zaret BL. Response of right ventricular ejection fraction to upright bicycle exercise in coronary artery disease. Circulation. 1979;60(6):1292–1300. doi:10.1161/01.cir.60.6.1292

50. Coma-Canella I, Del Val Gomez Martinez M, Terol I, Gallardo F, Castro Beiras J. Radionuclide assessment of right ventricular contractile reserve after acute myocardial infarction. Am J Cardiol. 1994;74(10):982–986. doi:10.1016/0002-9149(94)90844-3

51. Nakamura S, Iwasaka T, Kimura Y, et al. Right ventricular ejection fraction during exercise in patients with recent myocardial infarction: Effect of the interventricular septum. Am Heart J. 1994;127(1):49–55. doi:10.1016/0002-8703(94)90508-8

52. Karvounis HI, Papadopoulos CE, Ketikoglou DG, Zaglavara TA, Parharidis GE, Louridas GE. Usefulness of low-dose dobutamine stress echocardiography for the evaluation of spontaneous recovery of stunned myocardium in patients with acute right ventricular infarction. J Am Soc Echocardiogr Off Publ Am Soc Echocardiogr. 2005;18(4):351–356. doi:10.1016/j.echo.2004.11.003

53. Sade LE, Ozin B, Ulus T, et al. Right ventricular contractile reserve in mitral stenosis: implications on hemodynamic burden and clinical outcome. Int J Cardiol. 2009;135(2):193–201. doi:10.1016/j.ijcard.2008.03.050

54. Utsunomiya H, Hidaka T, Susawa H, et al. Exercise-Stress Echocardiography and Effort Intolerance in Asymptomatic/Minimally Symptomatic Patients With Degenerative Mitral Regurgitation Combined Invasive-Noninvasive Hemodynamic Monitoring. Circ Cardiovasc Imaging. 2018;11(9):e007282. doi:10.1161/CIRCIMAGING.117.007282

55. Vitel E, Galli E, Leclercq C, et al. Right ventricular exercise contractile reserve and outcomes after early surgery for primary mitral regurgitation. Heart Br Card Soc. 2018;104(10):855–860. doi:10.1136/heartjnl-2017-312097

56. Utsunomiya H, Izumi K, Tsuchiya A, et al. Role of anatomical regurgitant orifice area and right ventricular contractile reserve in severe tricuspid regurgitation. Eur Heart J Cardiovasc Imaging. 2022;23(7):989–1000. doi:10.1093/ehjci/jeac004

57. Ton VK, Ramani G, Hsu S, et al. High Right Ventricular Afterload Is Associated with Impaired Exercise Tolerance in Patients with Left Ventricular Assist Devices. ASAIO J Am Soc Artif Intern Organs 1992. 2021;67(1):39–45. doi:10.1097/MAT.0000000000001169

58. Tran T, Muralidhar A, Hunter K, et al. Right ventricular function and cardiopulmonary performance among patients with heart failure supported by durable mechanical circulatory support devices. J Heart Lung Transplant Off Publ Int Soc Heart Transplant. 2021;40(2):128–137. doi:10.1016/j.healun.2020.11.009

59. Takenaka S, Sato T, Nagai T, et al. Impact of right ventricular reserve on exercise capacity and quality of life in patients with left ventricular assist device. Am J Physiol Heart Circ Physiol. 2023;324(3):H355–H363. doi:10.1152/ajpheart.00626.2022

60. Agudo CA, Monivas Palomero V, Gonzalez Lopez E, Mingo Santos S. Prognostic value of exercise echocardiography in patients with wild-type transthyretin amyloidosis. Ups J Med Sci. 2022;127. doi:10.48101/ujms.v127.8410

61. Ladefoged B, Pedersen AD, Seefeldt J, et al. Exercise Hemodynamics and Mitochondrial Oxidative Capacity in Disease Stages of Wild-Type Transthyretin Amyloid Cardiomyopathy. J Am Heart Assoc. 2024;13(13):e034213. doi:10.1161/JAHA.124.034213

62. Berger HJ, Matthay RA, Davies RA. Comparison of exercise right ventricular performance in chronic obstructive pulmonary disease and coronary artery disease: Noninvasive assessment by quantitative radionuclide angiocardiography. Invest Radiol. 1979;14(5):342–353. doi:10.1097/00004424-197909000-00002

63. Rodriguez DA, Sancho-Munoz A, Rodo-Pin A, et al. Right Ventricular Response During Exercise in Patients with Chronic Obstructive Pulmonary Disease. Heart Lung Circ. 2017;26(6):631–634. doi:10.1016/j.hlc.2016.10.015

64. Rozenbaum Z, Ben-Gal Y, Kapusta L, et al. Combined Echocardiographic and Cardiopulmonary Exercise to Assess Determinants of Exercise Limitation in Chronic Obstructive Pulmonary Disease. J Am Soc Echocardiogr. 2021;34(2):146–155.e5. doi:10.1016/j.echo.2020.09.014

65. Martens P, Claessen G, Van De Bruaene A, et al. Iron Deficiency Is Associated With Impaired Biventricular Reserve and Reduced Exercise Capacity in Patients With Unexplained Dyspnea. J Card Fail. 2021;27(7):766–776. doi:10.1016/j.cardfail.2021.03.010

66. Sampaio F, Lamata P, Bettencourt N, et al. Assessment of cardiovascular physiology using dobutamine stress cardiovascular magnetic resonance reveals impaired contractile reserve in patients with cirrhotic cardiomyopathy. J Cardiovasc Magn Reson Off J Soc Cardiovasc Magn Reson. 2015;17:61. doi:10.1186/s12968-015-0157-6

67. Zimmermann P, Schierbauer J, Kopf N, et al. Speckle-Tracking Analysis of the Right and Left Heart after Peak Exercise in Healthy Subjects with Type 1 Diabetes: An Explorative Analysis of the AppEx Trial. J Cardiovasc Dev Dis. 2023;10(11). doi:10.3390/jcdd10110467

68. Chen Y, Gao Y, Zhou Y, et al. Human Immunodeficiency Virus Infection and Incident Heart Failure: A Meta-Analysis of Prospective Studies. JAIDS J Acquir Immune Defic Syndr. 2021;87(1):741–749. doi:10.1097/QAI.0000000000002629

69. Patterson AJ, Sarode A, Al-Kindi S, et al. Evaluation of dyspnea of unknown etiology in HIV patients with cardiopulmonary exercise testing and cardiovascular magnetic resonance imaging. J Cardiovasc Magn Reson Off J Soc Cardiovasc Magn Reson. 2020;22(1):74. doi:10.1186/s12968-020-00664-6

70. Xu B, Grimm RA, Jellis CL, et al. Teamwork using strain imaging in the echocardiographic assessment of right ventricular systolic function: A cardiac magnetic resonance imaging correlation study. Echocardiography. 2019;36(1):94–101. doi:10.1111/echo.14199

71. Collier P, Xu B, Kusunose K, et al. Impact of abnormal longitudinal rotation on the assessment of right ventricular systolic function in patients with severe pulmonary hypertension. J Thorac Dis. 2018;10(8):4696–4704. doi:10.21037/jtd.2018.07.118

72. Pergola V, Previtero M, Lorenzoni G, et al. Feasibility and role of right ventricular stress echocardiography in adult patients. J Cardiovasc Echography. 2021;31(2):68–72. doi:10.4103/jcecho.jcecho_4_21

73. Isaza N, Gonzalez M, Saijo Y, et al. Incremental Value of Global Longitudinal Strain to Michigan Risk Score and Pulmonary Artery Pulsatility Index in Predicting Right Ventricular Failure Following Left Ventricular Assist Devices. Heart Lung Circ. 2022;31(8):1110–1118. doi:10.1016/j.hlc.2022.03.012

74. Tello K, Dalmer A, Vanderpool R, et al. Right ventricular function correlates of right atrial strain in pulmonary hypertension: a combined cardiac magnetic resonance and conductance catheter study. Am J Physiol-Heart Circ Physiol. 2020;318(1):H156–H164. doi:10.1152/ajpheart.00485.2019

