## Supplementary material for "Right Ventricular Contractile Reserve: A Scoping Review": Online supplement - search strategy

Ovid MEDLINE(R) ALL <1946 to July 11, 2024>

1	((rv or right ventric*) and contractile reserve*).mp.	133

2	((rv or right ventric*) adj reserve*).mp.	40

3	1 or 2	163

Embase <1974 to 2024 July 11>

1	((rv or right ventric*) and contractile reserve*).mp.	337

2	((rv or right ventric*) adj reserve*).mp.	87

3	1 or 2	406

Search Name:	Siuba 7-12-24 rvcr

Date Run:	12/07/2024 22:43:33

Comment:

ID	Search	Hits

#1	(((rv or right NEXT ventric*) and contractile NEXT reserve*)):ti,ab,kw	11

#2	(((rv or right ventric*) NEXT reserve*)):ti,ab,kw	24

#3	#1 or #2	34

Cochrane Central Register of Controlled Trials

Issue 7 of 12, July 2024 = 34

Neil Nero, MLIS, Education Institute, Floyd D. Loop Alumni Library, Cleveland Clinic, Cleveland, OH, USA.

https://orcid.org/0000-0002-8391-4785
